# Implementation of a qPCR assay coupled with genomic surveillance for real-time monitoring of SARS-CoV-2 variants of concern

**DOI:** 10.1101/2021.05.20.21256969

**Authors:** Natalia Rego, Alicia Costábile, Mercedes Paz, Cecilia Salazar, Paula Perbolianachis, Lucía Spangenberg, Ignacio Ferrés, Rodrigo Arce, Alvaro Fajardo, Mailen Arleo, Tania Possi, Inés Bellini, Lucia Bilbao, Natalia Reyes, Ma Noel Bentancor, Andrés Lizasoain, María José Benítez, Matías Castells, Matías Victoria, Leticia Maya, Viviana Bortagaray, Ana Moller, Gonzalo Bello, Ighor Arantes, Mariana Brandes, Pablo Smircich, Odhille Chappos, Melissa Duquía, Belén González, Luciana Griffero, Mauricio Méndez, Ma Pía Techera, Juan Zanetti, Bernardina Rivera, Matías Maidana, Martina Alonso, Cecilia Alonso, Julio Medina, Henry Albornoz, Rodney Colina, Veronica Noya, Gregorio Iraola, Tamara Fernández-Calero, Gonzalo Moratorio, Pilar Moreno

## Abstract

We developed a genomic surveillance program for real-time monitoring of SARS-CoV-2 variants of concern in Uruguay. Here, we present the first results, including the proposed qPCR-VOC method, the general workflow and the report of the introduction and community transmission of the VOC P.1 in Uruguay in multiple independent events.

## Introduction

Throughout the pandemic, as part of the natural viral evolution, genetic variants of SARS-CoV-2 have emerged. Some of these have shown increased transmissibility, more severe disease with increased hospitalizations, intensive care units admissions and deaths, significant reduction in neutralization by antibodies generated during previous infection or vaccination, reduced effectiveness of treatments or vaccines and/or diagnostic detection failures [1, 2]. These are called variants of concern (VOCs). To date, five VOCs have been identified: B.1.1.7, B.1.351, P.1, B.1.427 and B.1.429 [3]. A robust surveillance workflow for early identification of VOCs is key to accelerate pandemic responsiveness.

Surveillance data from Brazil demonstrated a sharp increase in the number of SARS-CoV-2 cases, hospitalisations and deaths after the emergence of the VOC P.1 in the Amazonas state in November 2020 [4, 5]. The VOC P.1 displays a higher transmissibility than previous local SARS-CoV-2 lineages [4, 5] and rapidly spread through the country, becoming the predominant strain in most country regions by February-March, 2021 [6]. This VOC has also spread worldwide and as of 4 May 2021, the lineage P.1 has been detected in at least 41 countries worldwide [7]. This raises the concern that VOC P.1 might follow in other countries the same epidemic trajectory observed in Brazil.

Uruguay, which shares 600 miles of dry border with Brazil and hosts more than 170,000 people living in twin cities with intense economic and social interaction, has experienced an exponential increase of COVID-19 cases since late February, 2021, placing it at the top of the countries with the higher number of daily confirmed cases and deaths per million people. Despite the Brazilian-Uruguayan border has remained closed to nonresident foreigners since March 2020, there is evidence of high viral flux between both countries [8, 9], implying the risk for imminent introduction of P.1 into Uruguay and the necessity of coming up with an organized strategy to monitorize this and other viral variants circulating in the country.

We developed a comprehensive genomic surveillance program for real-time monitoring VOCs emergence in Uruguay to help sanitary authorities to manage the crisis. We created an inter-institutional working group (IiWG) to run this program aiming to put together: i) a diagnostic network, ii) expertise and resources to handle large-scale sequencing, iii) computational scientists to analyze genomic datasets, and iv) an affordable and decentralizable “in house” qPCR test specifically designed to detect worldwide known VOCs (B.1.1.7, B.1.351 and P.1). Here, we present the results of the first two weeks of the IiWG, including the proposed qPCR-VOC method, the general workflow and the report of the introduction and community transmission of the VOC P.1 in Uruguay in multiple independent events.

## Results

### Inter-institutional working group (IiWG) for real time genomic surveillance of SARS-CoV-2

This task force is form by four Institutions (Institut Pasteur de Montevideo, Universidad de la República, Zurgen-Sanatorio Americano and Ministerio de Salud Pública), including more than 30 people, among them researchers, physicians and health authorities. Four diagnostic laboratories are included in the network, that are able to process all together more than 3,000 nasopharyngeal samples per day. Of those, between 200 – 300 SARS-CoV2 positive samples are received weekly for qPCR-VOC analysis and 50% of them were further processed for SARS-CoV-2 genome sequencing. Those numbers are projected to increase with time if exponential growth of cases is sustained. Figure 1A shows the IiWG workflow.

**Figure 1.**
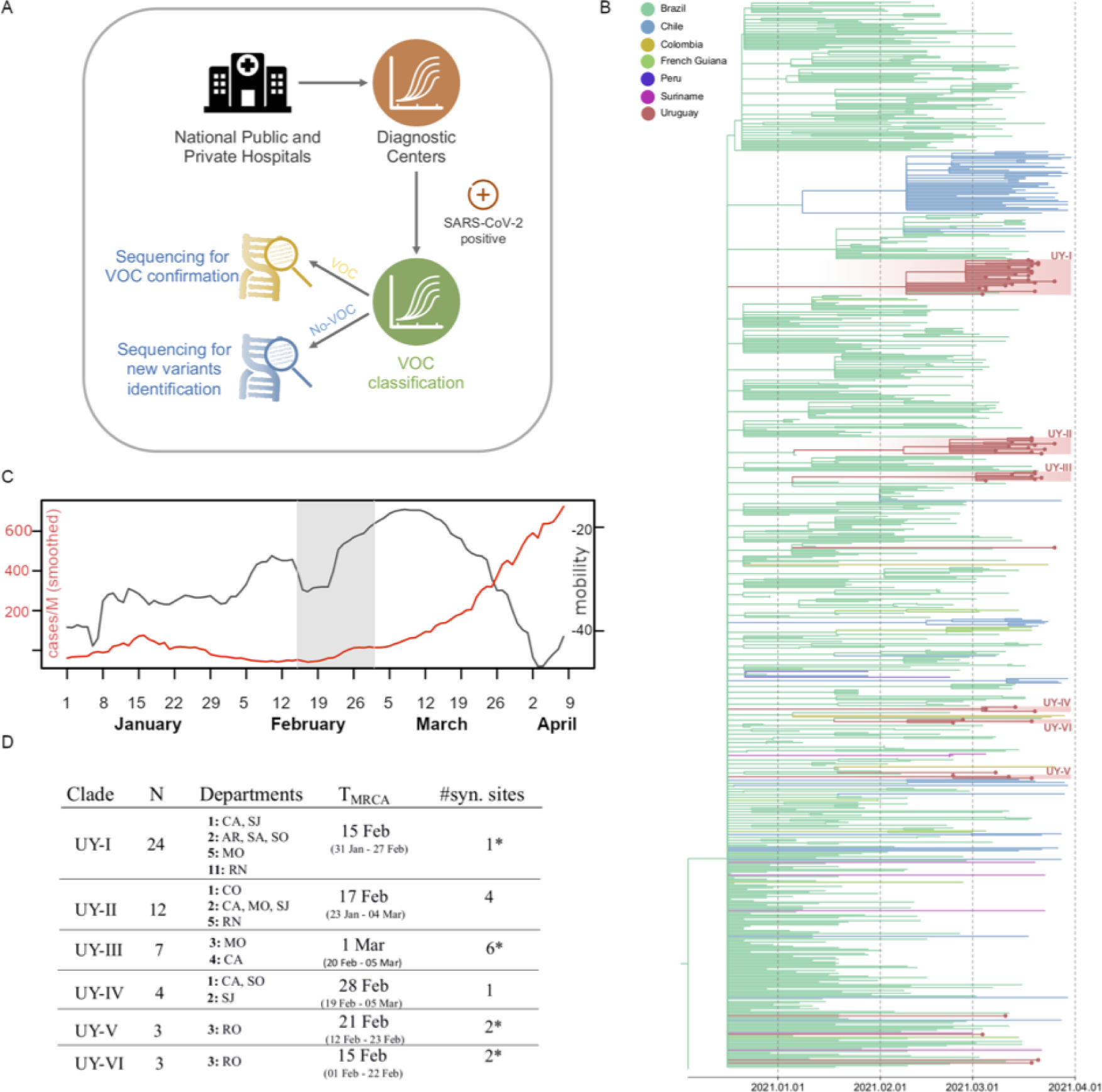
1A. Inter-institutional working group (IiWG) Workflow. Diagnostic centers perform standard qPCR for identification of positive samples. Those are further evaluated by a drop-out qPCR assay to determine known VOCs. A subset of samples classified as VOC and NO-VOC by drop-out qPCR are then sequenced to confirm known VOCs and detect new potential variants. 1B. Time-scaled Bayesian phylogeographic MCC tree of 59 Uruguayan and 691 South American P.1 whole genome sequences. The tree was rooted with the EPI_ISL_833137 sequence (collection date 2020-12-04). Branches are colored according to the most probable location state of their descendant nodes as indicated at the legend. As can be seen from the read blocks distribution along the P.1 tree, at least 12 independent introductions can be estimated, with locally transmitted clusters of size three to 24. Brazil, in aquamarine, has been the source of dissemination of P.1 sequences to Uruguay and other South American countries. 1.C Mobility index (black) and number of cases per million (red) is shown daily. Mobility index is calculated as in https://hdl.handle.net/20.500.12008/27166. The time ranges from January 1st to 9th April. In gray the estimated time period where the P.1 was most likely first introduced to the country (∼15^th^ February). 1.D Summary table with the six (non-singleton) clades found in the time-scaled Bayesian tree. Clades are numbered from UY-I to UY-VI. N represents the number of sequences in each clade. The Department column holds information of the collection sites and number of sequences per site. CA:Canelones, SJ:San Jose, MO:Montevideo, RO:Rocha, SA:Salto, CO:Colonia, RN: Rio Negro, AR:Artigas, SO: Soriano. T_MRCA_ (most recent common ancestor) is the estimated time of the clade (in brackets the boundaries of the 95% confidence interval). The syn. sites columns corresponds to the number of synapomorphic sites defining each clade. Additionally, the asterisk marks whether the amino acid replacement is considered radical. See Supplementary Table 3 for more details on the clades and synapomorphies.

### Detection of P.1 variant by VOCs qPCR method and sequencing

In the first two weeks of the IiWG work, a total of 251 SARS-CoV-2 positive RNA samples were obtained from hospitalised cases and outpatient cases collected from all over the country (15 out of 19 Uruguayan departments) between 01/11/2021 and 03/26/2021 (Supplementary Table 1) and processed according to Figure 1A. Patients involved in the study presented Cts for initial diagnostic PCR which ranged from 9 to 34.7, both genders were included (95 males, 95 females and 61 unknown) and age ranged from 1 to 85 years old.

Results from qPCR-VOC assay showed that 67/251 samples (27%) were classified as putative P.1/B.1.351 (Supplementary Figure 1). All VOC positives samples, plus 31 additional samples, were sequenced using the ARCTIC Network protocol for the MinION platform (Oxford Nanopore)[10] as described in Supplementary Methods, to validate PCR-VOC classification. For 74 final consensus sequences with high quality that were assigned to SARS-CoV-2 lineages using Pango nomenclature [11], we achieved a 100% agreement between qPCR-VOC and genome sequencing results. As expected, given the proximity to Brazil, samples classified as P.1/B.1.351 by qPCR-VOC, were assigned to lineage P.1 after genome sequencing. qPCR-VOC is a feasible, precise and scalable method to follow known VOCs in real-time.

Given the qPCR-VOC screening and the confirmation of P.1 by genome sequencing, we established the circulation of lineage P.1 in 15 of 19 departments in Uruguay here analyzed (Figure 1D and Supplementary Figure 1 and 2). To estimate the geographic source and the number of independent introductions of P.1 variant into Uruguay, P.1 Uruguayan sequences obtained in this work were combined with other P.1 sequences from Uruguay (n=3) collected in February (Supplementary Table 1), and with 691 P.1 sequences from South America available in EpiCoV/GISAID [12] (Supplementary Table 2). The ML phylogeographic analysis identified at least 12 independent introductions of lineage P.1 in Uruguay from Brazil and at least six local transmission clusters (designated as UY-I to UY-VI) of between three and 24 sequences (Figure 1B). The median T_MRCA_ of these six Uruguayan P.1 clades was estimated with Bayesian analysis to between mid-February and early March 2021 (Figure 1D, Supplementary Figure 3 and Supplementary Table 3), which coincides with a period of increasing mobility in the country and the beginning of the exponential growth phase (Figure 1C).

## Conclusion

The rapid emergence of the SARS-CoV-2 lineage P.1 in the South American region requires the need for increased screening of this highly transmissible virus. Here we developed a comprehensive genomic surveillance program and provide a clear example of how multidisciplinary teams are key mechanisms for helping sanitary authorities to better manage the crisis. Thus, we created an inter-institutional working group with diagnostic laboratories, hospitals (public and private), public and academic institutions across Uruguay that allows real-time VOCs surveillance. Our findings revealed that the VOC P.1 was introduced in Uruguay at multiple times over a period of increasing mobility between mid-February and early March, 2021. The introduction of the highly transmissible VOC P.1 coupled to the increasing human mobility probably contributed to the rapid local spread of this variant and to the COVID-19 epidemic worsening in Uruguay in 2021.

## Data Availability

Data was uploaded to GISAID

## Acknowledgements

Thanks for efforts from different groups that contribute with SARS-CoV-2 genomes to the EpiCoV GISAID initiative (Supplementary GISAID Acknowledgment Table). We thank Marcelo Fiori and María Inés Fariello for sharing the Uruguayan mobility data and Christian Brandt from Institute for Infectious Diseases and Infection Control (Jena University Hospital), for adapting their poreCov Nextflow pipeline to our requirements and to quickly clarify doubts about its implementation.

We also want to thank Dr. Nicolas Nin and Javier Hurtado from Hospital Español.

## Supplementary Material File

Supplementary Methods

Supplementary References

Supplementary Tables

Supplementary Figures

Supplementary GISAID

Acknowledgment Table

## 1. Supplementary Methods

### Ethics statement

Residual de-identified RNA samples from positive patients were remitted to the Institut Pasteur Montevideo (IPmon). IPmon was validated by the Ministry of Health of Uruguay as an approved center providing diagnostic testing for COVID-19. All samples were de-identified before receipt by the study investigators.

### Development of a “drop out” qPCR SARS-CoV-2 approach to detect VOCs

Primers and “drop out” probes designed to target the deletion sites in both Spike (S) and ORF1ab viral genes were used for this assay. Primers and probes targeting ORF1ab deletion (Δ3675-3677SGF) were designed using the nucleotide sequence from Wuhan strain (GenBank acc. NC_045512) and are shown in Supplementary Table 4. Oligonucleotide complementarity and Tm were assessed with OligoAnalyzer (IDT)[1]. Primers and probes targeting S deletion (Δ69/70 HV) were taken from [2]. B.1.1.7 has both deletions whereas P.1 and B.1.351 have only deletion in ORF1ab.

Hence, we expect no amplification with both probes with B.1.1.7 lineage viruses, only amplification of the S gene with P.1 and B.1.351 lineages and amplification of both genomic regions with other lineages. These primer/probe sets were analyzed with BLAST to rule out similarities with other sequences other than SARS-CoV-2. The detection strategy is based on the standard hydrolysis probe system (TaqMan® Technology). The S gene specific probes are labeled with the HEX fluorophore, while the Orf1a probe is custom labeled with CY5 fluorophore. The primers and probes were synthesized by Integrated DNA Technologies (IDT) and purified by high-pressure liquid chromatography. The OneStep Reaction mix was prepared using 5 µl of RNA samples, 5 µl of 4 x TaqMan Fast Virus 1-Step Master Mix (Thermo Cat. No. 4444436), with 0.3 µM of each S primers, 0.1 µM of S probe, 0.3 µM of each Orf1a primer, 0.1 µM of Orf1a probe and molecular biology grade water to a final volume of 20 µl. The RT-qPCR assays were performed using QuantStudio™ 7 (Applied Biosystem).

### Genome Sequencing

Sequencing libraries were prepared according to the Eco PCR tiling of COVID-19 virus protocol (Oxford Nanopore Technologies, United Kingdom), based on the method described by Quick J. [3, 4] with some modifications. RNA samples previously screened by a qPCR assay, were reverse transcribed using SuperScript™ II Reverse Transcriptase (Thermo Fisher Scientific Inc., MA, USA) or the LunaScript® RT SuperMix Kit (New England Biolabs, Ipswich, MA, USA). A negative control was included at this point and carried throughout the protocol. V3 Artic Network primers (IDT) were used for SARS-CoV-2 genome amplification using the Q5® High-Fidelity DNA Polymerase (NEB) in two multiplex PCR reactions. At this point a positive control from a previously sequenced sample was added and carried throughout the protocol. Both reactions were pooled and diluted for the end prep reaction using the NEBNext Ultra II End repair / dA-tailing Module (NEB). Blunt/TA Ligase master mix (NEB) was used to ligate unique barcodes (Native Barcoding expansion kit, EXP-NBD196, Oxford Nanopore Technologies, United Kingdom) to the end prepped samples. Barcoded samples were pooled, cleaned up using 0.4X volume of AMPure XP beads (Beckman coulter, USA) and quantified using the Qubit HS dsDNA kit (Thermo Fisher Scientific Inc., MA, USA). AMII sequencing adapters were ligated using the NEBNext Quick Ligation Module (New England Biolabs, Ipswich, MA, USA) and washed with SFB buffer (Oxford Nanopore Technologies, United Kingdom). Final library was eluted on EB buffer (Oxford Nanopore Technologies, United Kingdom) and quantified using the Qubit HS dsDNA kit. Approximately 50 fmol was loaded into a FLO-MIN106D R9.4.1 flow cell and sequenced on the GridION X5 sequencing platform (ONT) until a minimum sequencing depth of 500X was achieved.

Basecalling and demultiplexing was performed with Guppy 4.3.2 [5] using the high accuracy mode. Samples were demultiplexed using both front and rear barcodes using the --requiere_both_barcodes option. Consensus genomes were generated and using the poreCov pipeline [6, 7] implemented in Nextflow [8], which includes the Artic Network workflow [9], Pangolin for PANGO lineage [10] assignment, nextclade for clade assignment [11], Kraken2 and Krona for contamination detection and visualization [12, 13], minimap2 for read mapping [14], medaka for consensus generation, and using singularity as container engine [15]. Amplicons that are not sequenced or whose depth is less than 20x are not included in the consensus sequences and these positions are represented by “N” stretches. Complete sequences with up to 15% of Ns were kept for further analysis. All genomes obtained in this study were uploaded to the EpiCoV database in the GISAID initiative under the accession numbers: EPI_ISL_1992241, EPI_ISL_1992100, EPI_ISL_1991974, and EPI_ISL_2031707 to EPI_ISL_2031762 and additional metadata for Uruguayan genomes is presented in Supplementary Table 1.

### Phylogenetic and phylogeographic analysis

Uruguayan P.1 genome sequences (n = 59) were manually curated in specific genome positions of interest, such as P.1 synapomorphies, and were analyzed in the context of additional P.1 sequences from South America, downloaded from EpiCoV database in the GISAID initiative [16]. Downloaded sequences (n = 905, see Supplementary GISAID Acknowledgment Table) were complete, high quality, with full collection date information and sampled before March 31, 2021. Alignment was performed with MAFFT v7.471 [17]. After carefully alignment inspection, several sequences were missing the characteristic indels (del11288-11296 and ins28269-28273) of P.1 and were excluded, retaining 691 sequences from Brazil (n = 590), Chile (n = 72), Colombia (n = 4), French Guiana (n = 15), Peru (n = 2) and Suriname (n = 8) (Supplementary Table 2). Maximum likelihood (ML) phylogenetic analysis of P.1 samples was conducted using IQ-TREE version 1.6.12 under the GTR+F+R3 nucleotide substitution model selected by the built-in ModelFinder option [18]. Branch support was assessed by the approximate likelihood-ratio test based on a Shimodaira-Hasegawa-like procedure with 1,000 replicates. The tree was rooted using the sequence EPI_ISL_833137 with collection date 2020-12-04 and visualized using iTOL [19]. ML phylogeographic analysis and time-scaled Bayesian analysis were next performed to infer the geographical source of Uruguayan P.1 samples and to estimate the time of the most recent common ancestors (T_MRCA_) of the Uruguayan P.1 clades, respectively. The previously generated ML phylogenetic tree was time-scaled using TreeTime [20] applying a fixed clock rate of 8 x 10-4 substitutions/site/year, based on previous estimates [21, 22], a skyline coalescent model with eight grid points and keeping polytomies. The time-scaled tree was then employed for the ancestral character state reconstruction (ACR) of epidemic locations with PastML [23], using the Marginal Posterior Probabilities Approximation (MPPA) method with an F81-like model. A time-scaled Bayesian phylogenetic tree was constructed using the Bayesian Markov Chain Monte Carlo (MCMC) approach implemented in BEAST 1.10 [24] with BEAGLE library v3 to improve computational time. Bayesian analysis was conducted using: the non-parametric Bayesian skyline (BSKL) model as the coalescent tree prior [25], the GTR+I+G model of nucleotide substitution and a strict molecular clock model with a uniform prior on substitution rate (8-10 x 10-4 substitutions/site/year). Two MCMC chains were run for 50 million generations and then combined to ensure stationarity and good mixing. Convergence (Effective Sample Size > 200) in parameter estimates was assessed using TRACER v1.7 [26]. The maximum clade credibility (MCC) tree was summarized with TreeAnnotator v1.10 [27] and visualized using FigTree v1.4.4 [28].

## 3. Supplementary Tables

**Supplementary Table 1.** Information on the 251 analyzed samples. ID column, Ct, Department (collection site in Uruguay), and the result of the VOC-qPCR. Possible outputs in this case are either no-VOC or P.1/B.1.351. The asterisk marks those samples with successful sequencing. The ID identifiers do not reveal the identity of the study subjects.

| ID | Ct | Department | Variants by PCR |
| --- | --- | --- | --- |
| CUY1-000001 | 20.1 | Treinta y Tres | No-VOC |
| CUY1-000002 | 23.8 | Treinta y Tres | No-VOC |
| CUY1-000004 | 28.7 | Artigas | No-VOC |
| CUY1-000006 | 18.0 | Artigas | No-VOC |
| CUY1-000007 | 21.5 | San José | No-VOC |
| CUY1-000008 | 22.7 | San José | No-VOC |
| CUY1-000011 | 22.8 | Artigas | P.1/B.1.351* |
| CUY1-000012 | 23.0 | Artigas | No-VOC |
| CUY1-000015 | 23.5 | Artigas | No-VOC |
| CUY1-000017 | 29.7 | Montevideo | No-VOC |
| CUY1-000020 | 34.7 | Montevideo | P.1/B.1.351* |
| CUY1-000021 | 22.9 | Colonia | No-VOC |
| CUY1-000022 | 19.6 | Maldonado | No-VOC |
| CUY1-000023 | 18.3 | Maldonado | No-VOC |
| CUY1-000024 | 26.9 | Montevideo | No-VOC |
| CUY1-000025 | 15.4 | Montevideo | P.1/B.1.351* |
| CUY1-000028 | 31.5 | Salto | No-VOC |
| CUY1-000029 | 18.7 | Salto | No-VOC |
| CUY1-000030 | 28.7 | Colonia | NC |
| CUY1-000031 | 21.3 | Artigas | P.1/B.1.351* |
| CUY1-000032 | 22.9 | San José | No-VOC |
| CUY1-000034 | 29.5 | Canelones | No-VOC |
| CUY1-000035 | 27.9 | Canelones | No-VOC |
| CUY1-000036 | 29.1 | Salto | No-VOC |
| CUY1-000037 | 29.9 | Flores | No-VOC |
| CUY1-000038 | 25.4 | Salto | No-VOC |
| CUY1-000039 | 24.4 | Flores | No-VOC |
| CUY1-000040 | 27.9 | Tacuarembó | No-VOC |
| CUY1-000041 | 30.1 | Flores | No-VOC |
| CUY1-000042 | 19.0 | Canelones | No-VOC |
| CUY1-000043 | 29.6 | Canelones | No-VOC |
| CUY1-000045 | 20.8 | ND | P.1/B.1.351* |
| CUY1-000046 | 25.9 | Canelones | P.1/B.1.351* |
| CUY1-000047 | 25.5 | Canelones | P.1/B.1.351* |
| CUY1-000048 | 22.4 | Salto | No-VOC |
| CUY1-000049 | 20.2 | San José | P.1/B.1.351* |
| CUY1-000050 | 22.7 | San José | P.1/B.1.351* |
| CUY1-000051 | 24.9 | Salto | No-VOC |
| CUY1-000052 | 25.2 | Salto | No-VOC |
| CUY1-000053 | 33.0 | Tacuarembó | No-VOC |
| CUY1-000054 | 27.5 | Salto | No-VOC |
| CUY1-000055 | 20.9 | Salto | No-VOC |
| CUY1-000056 | 18.7 | Canelones | No-VOC |
| CUY1-000057 | 21.8 | Tacuarembó | No-VOC |
| CUY1-000060 | 22.8 | Florida | No-VOC |
| CUY1-000061 | 16.4 | Rocha | No-VOC |
| CUY1-000062 | 18.2 | Rocha | P.1/B.1.351* |
| CUY1-000063 | 20.0 | Florida | No-VOC |
| CUY1-000064 | 21.9 | Río Negro | No-VOC |
| CUY1-000065 | 20.2 | Rocha | No-VOC |
| CUY1-000066 | 22.4 | Rocha | No-VOC |
| CUY1-000067 | 22.1 | Rocha | No-VOC |
| CUY1-000068 | 31.8 | Flores | No-VOC |
| CUY1-000069 | 25.5 | Flores | No-VOC |
| CUY1-000070 | 30.5 | Flores | P.1/B.1.351 |
| CUY1-000071 | 34.1 | Río Negro | P.1/B.1.351 |
| CUY1-000072 | 25.9 | Río Negro | P.1/B.1.351* |
| CUY1-000073 | 22.2 | Flores | No-VOC |
| CUY1-000074 | ND | ND | No-VOC |
| CUY1-000075 | 16.4 | San José | P.1/B.1.351* |
| CUY1-000076 | 19.2 | Canelones | No-VOC |
| CUY1-000077 | 20.9 | San José | No-VOC |
| CUY1-000078 | 26.8 | Salto | No-VOC |
| CUY1-000079 | 26.3 | Salto | No-VOC |
| CUY1-000080 | 19.9 | Río Negro | P.1/B.1.351* |
| CUY1-000081 | 29.5 | Artigas | P.1/B.1.351 |
| CUY1-000082 | 19.2 | Artigas | No-VOC |
| CUY1-000083 | 17.2 | Artigas | No-VOC |
| CUY1-000084 | 25.5 | ND | P.1/B.1.351 |
| CUY1-000085 | 18.1 | Salto | No-VOC |
| CUY1-000086 | 19.4 | Colonia | No-VOC |
| CUY1-000087 | 26.6 | Florida | No-VOC |
| CUY1-000088 | 23.9 | Flores | No-VOC |
| CUY1-000089 | 18.6 | San José | No-VOC |
| CUY1-000090 | 21.4 | San José | P.1/B.1.351* |
| CUY1-000091 | 22.2 | Montevideo | No-VOC |
| CUY1-000092 | 20.1 | Canelones | No-VOC |
| CUY1-000093 | 19.0 | Canelones | P.1/B.1.351* |
| CUY1-000094 | 17.6 | Canelones | P.1/B.1.351* |
| CUY1-000095 | 23.0 | San José | P.1/B.1.351* |
| CUY1-000096 | 19.9 | Montevideo | No-VOC |
| CUY1-000097 | 29.2 | Montevideo | P.1/B.1.351* |
| CUY1-000098 | 24.5 | ND | No-VOC |
| CUY1-000099 | 29.7 | Canelones | No-VOC |
| CUY1-000100 | 15.0 | Salto | No-VOC |
| CUY1-000101 | 11.0 | Salto | No-VOC |
| CUY1-000102 | 9.0 | Salto | No-VOC |
| CUY1-000103 | 13.0 | Salto | No-VOC |
| CUY1-000104 | 15.0 | Salto | No-VOC |
| CUY1-000105 | 11.0 | Salto | No-VOC |
| CUY1-000106 | 12.0 | Salto | No-VOC |
| CUY1-000107 | 15.0 | Río Negro | No-VOC |
| CUY1-000108 | 12.0 | Salto | No-VOC |
| CUY1-000109 | 12.0 | Salto | No-VOC |
| CUY1-000110 | 13.0 | Río Negro | P.1/B.1.351* |
| CUY1-000111 | 13.0 | Salto | No-VOC |
| CUY1-000112 | 14.0 | Salto | No-VOC |
| CUY1-000113 | 12.0 | Salto | No-VOC |
| CUY1-000114 | 10.0 | Río Negro | P.1/B.1.351* |
| CUY1-000115 | 17.0 | Río Negro | P.1/B.1.351* |
| CUY1-000116 | 14.0 | Salto | No-VOC |
| CUY1-000117 | 14.0 | Salto | No-VOC |
| CUY1-000118 | 17.0 | Salto | No-VOC |
| CUY1-000119 | 10.0 | Salto | No-VOC |
| CUY1-000120 | 18.0 | Salto | No-VOC |
| CUY1-000121 | 12.0 | Salto | No-VOC |
| CUY1-000122 | 13.0 | Salto | No-VOC |
| CUY1-000123 | 12.0 | Salto | No-VOC |
| CUY1-000124 | 17.0 | Salto | No-VOC |
| CUY1-000125 | 13.0 | Salto | P.1/B.1.351* |
| CUY1-000126 | 16.0 | Salto | No-VOC |
| CUY1-000127 | 18.0 | Salto | No-VOC |
| CUY1-000128 | 16.0 | Salto | No-VOC |
| CUY1-000129 | 11.0 | Salto | P.1/B.1.351* |
| CUY1-000130 | 18.0 | Tacuarembó | No-VOC |
| CUY1-000131 | 15.0 | Tacuarembó | No-VOC |
| CUY1-000132 | 17.0 | Tacuarembó | No-VOC |
| CUY1-000133 | 19.0 | Tacuarembó | No-VOC |
| CUY1-000134 | 17.0 | Tacuarembó | No-VOC |
| CUY1-000135 | 16.0 | Tacuarembó | No-VOC |
| CUY1-000136 | 19.0 | Tacuarembó | No-VOC |
| CUY1-000137 | 15.0 | Tacuarembó | No-VOC |
| CUY1-000138 | 18.0 | Tacuarembó | No-VOC |
| CUY1-000139 | 14.0 | Tacuarembó | No-VOC |
| CUY1-000140 | 19.0 | Tacuarembó | No-VOC |
| CUY1-000141 | 13.0 | Tacuarembó | No-VOC |
| CUY1-000142 | 19.0 | Tacuarembó | No-VOC |
| CUY1-000143 | 18.6 | Tacuarembó | No-VOC |
| CUY1-000144 | 18.0 | Tacuarembó | No-VOC |
| CUY1-000145 | 18.2 | Tacuarembó | No-VOC |
| CUY1-000146 | 14.2 | Tacuarembó | No-VOC |
| CUY1-000147 | 15.7 | Tacuarembó | No-VOC |
| CUY1-000148 | 18.0 | Tacuarembó | No-VOC |
| CUY1-000149 | 18.5 | Tacuarembó | No-VOC |
| CUY1-000150 | 15.2 | Tacuarembó | No-VOC |
| CUY1-000151 | 18.0 | Tacuarembó | No-VOC |
| CUY1-000152 | 16.3 | Tacuarembó | No-VOC |
| CUY1-000153 | 18.5 | Tacuarembó | No-VOC |
| CUY1-000154 | 18.5 | Tacuarembó | No-VOC |
| CUY1-000155 | 19.2 | Tacuarembó | No-VOC |
| CUY1-000156 | 18.1 | Tacuarembó | No-VOC |
| CUY1-000157 | 19.5 | Tacuarembó | No-VOC |
| CUY1-000158 | 19.8 | Tacuarembó | No-VOC |
| CUY1-000159 | 18.7 | Tacuarembó | No-VOC |
| CUY1-000160 | 19.4 | Tacuarembó | No-VOC |
| CUY1-000161 | 15.2 | Tacuarembó | No-VOC |
| CUY1-000162 | 11.8 | Tacuarembó | No-VOC |
| CUY1-000163 | 17.5 | Tacuarembó | No-VOC |
| CUY1-000164 | 17.3 | Tacuarembó | No-VOC |
| CUY1-000165 | 14.1 | Tacuarembó | No-VOC |
| CUY1-000166 | 17.3 | Tacuarembó | No-VOC |
| CUY1-000167 | 15.3 | Tacuarembó | No-VOC |
| CUY1-000168 | 15.3 | Tacuarembó | No-VOC |
| CUY1-000169 | 14.5 | Tacuarembó | No-VOC |
| CUY1-000170 | 15.5 | Tacuarembó | No-VOC |
| CUY1-000171 | 13.2 | Tacuarembó | No-VOC |
| CUY1-000172 | 13.4 | Tacuarembó | No-VOC |
| CUY1-000173 | 17.9 | Tacuarembó | No-VOC |
| CUY1-000174 | 16.5 | Tacuarembó | No-VOC |
| CUY1-000175 | 17.3 | Tacuarembó | No-VOC |
| CUY2-000176 | 14.0 | Paysandú | No-VOC |
| CUY2-000177 | 13.0 | Paysandú | No-VOC |
| CUY2-000178 | 12.0 | Salto | No-VOC |
| CUY2-000179 | 14.0 | Salto | No-VOC |
| CUY2-000180 | 17.0 | Paysandú | No-VOC |
| CUY2-000181 | 16.0 | Paysandú | No-VOC |
| CUY2-000182 | 14.0 | Salto | No-VOC |
| CUY2-000183 | 12.0 | Montevideo | No-VOC |
| CUY2-000184 | 16.0 | Salto | No-VOC |
| CUY2-000185 | 17.0 | Salto | No-VOC |
| CUY2-000186 | 9.0 | Salto | No-VOC |
| CUY2-000187 | 9.0 | Salto | No-VOC |
| CUY2-000188 | 12.0 | Salto | No-VOC |
| CUY2-000189 | 14.0 | Salto | No-VOC |
| CUY2-000190 | 13.0 | Salto | No-VOC |
| CUY2-000191 | 16.0 | Paysandú | No-VOC |
| CUY2-000192 | 16.0 | Paysandú | No-VOC |
| CUY2-000193 | 14.0 | Paysandú | No-VOC |
| CUY2-000194 | 16.0 | Paysandú | No-VOC |
| CUY2-000195 | 11.0 | Salto | No-VOC |
| CUY2-000196 | 12.0 | Salto | No-VOC |
| CUY2-000197 | 11.0 | Salto | No-VOC |
| CUY2-000198 | 13.0 | Salto | P.1/B.1.351* |
| CUY2-000199 | 14.0 | Río Negro | P.1/B.1.351* |
| CUY2-000200 | 12.0 | Río Negro | P.1/B.1.351* |
| CUY2-000201 | 22.9 | Montevideo | No-VOC |
| CUY2-000202 | 20.0 | Montevideo | No-VOC |
| CUY2-000203 | 16.8 | Montevideo | P.1/B.1.351* |
| CUY2-000204 | 13.6 | Montevideo | P.1/B.1.351* |
| CUY2-000205 | 24.5 | Montevideo | No-VOC |
| CUY2-000206 | 13.5 | Montevideo | No-VOC |
| CUY2-000207 | 28.2 | Montevideo | No-VOC |
| CUY2-000208 | 22.7 | Tacuarembó | No-VOC |
| CUY2-000209 | 21.8 | Tacuarembó | No-VOC |
| CUY2-000210 | 31.5 | Artigas | No-VOC |
| CUY2-000211 | 22.3 | Río Negro | P.1/B.1.351* |
| CUY2-000212 | 21.9 | Río Negro | P.1/B.1.351* |
| CUY2-000213 | 20.3 | Río Negro | P.1/B.1.351* |
| CUY2-000214 | 21.9 | Río Negro | P.1/B.1.351* |
| CUY2-000215 | 23.6 | Artigas | P.1/B.1.351* |
| CUY2-000216 | 28.8 | Soriano | P.1/B.1.351 |
| CUY2-000217 | 21.0 | San José | No-VOC |
| CUY2-000218 | 24.2 | Soriano | No-VOC |
| CUY2-000219 | 22.7 | Soriano | No-VOC |
| CUY2-000220 | ND | ND | No-VOC |
| CUY2-000221 | 24.4 | Rocha | No-VOC |
| CUY2-000222 | 22.8 | San José | P.1/B.1.351* |
| CUY2-000223 | 25.9 | San José | No-VOC |
| CUY2-000224 | 23.2 | Maldonado | No-VOC |
| CUY2-000225 | 23.4 | Canelones | P.1/B.1.351 |
| CUY2-000226 | 22.0 | Canelones | No-VOC |
| CUY2-000227 | 23.0 | Rocha | No-VOC |
| CUY2-000228 | 19.7 | Rocha | P.1/B.1.351* |
| CUY2-000229 | 20.0 | Rocha | No-VOC |
| CUY2-000231 | 18.1 | Canelones | P.1/B.1.351* |
| CUY2-000232 | 24.4 | Soriano | No-VOC |
| CUY2-000233 | 21.3 | Rocha | P.1/B.1.351* |
| CUY2-000234 | 20.5 | Canelones | No-VOC |
| CUY2-000235 | 19.9 | Río Negro | P.1/B.1.351* |
| CUY2-000236 | 19.2 | Río Negro | P.1/B.1.351* |
| CUY2-000237 | 19.6 | Río Negro | P.1/B.1.351* |
| CUY2-000238 | 23.6 | Río Negro | P.1/B.1.351* |
| CUY2-000239 | 26.2 | Río Negro | P.1/B.1.351* |
| CUY2-000240 | 12.3 | Río Negro | P.1/B.1.351* |
| CUY2-000241 | 22.8 | Canelones | No-VOC |
| CUY2-000242 | 19.5 | Canelones | No-VOC |
| CUY2-000243 | 24.7 | Canelones | No-VOC |
| CUY2-000244 | 25.9 | Canelones | P.1/B.1.351* |
| CUY2-000245 | 18.0 | Canelones | P.1/B.1.351* |
| CUY2-000246 | 30.3 | Canelones | P.1/B.1.351 |
| CUY2-000247 | 21.5 | San José | No-VOC |
| CUY2-000249 | 23.5 | Canelones | No-VOC |
| CUY2-000250 | 21.6 | Artigas | P.1/B.1.351* |
| CUY2-000251 | 31.2 | Montevideo | No-VOC |
| CUY2-000252 | 17.2 | Montevideo | P.1/B.1.351* |
| CUY2-000253 | 23.3 | Montevideo | No-VOC |
| CUY2-000254 | 20.6 | Montevideo | P.1/B.1.351* |
| CUY2-000255 | 24.6 | Montevideo | No-VOC |
| CUY2-000256 | 19.5 | Montevideo | P.1/B.1.351* |
| CUY2-000257 | 23.2 | Canelones | P.1/B.1.351* |
| CUY2-000258 | 17.7 | Montevideo | P.1/B.1.351* |
| CUY2-000259 | 26.8 | Soriano | P.1/B.1.351* |
| CUY2-000260 | 25.3 | Soriano | P.1/B.1.351* |
| CUY2-000261 | 26.7 | Colonia | P.1/B.1.351* |
| CUY2-000262 | 22.7 | Montevideo | P.1/B.1.351 |
| CUY2-000263 | 18.3 | Montevideo | P.1/B.1.351* |
| CUY2-000264 | 23.6 | Montevideo | P.1/B.1.351 |
| CUY2-000265 | 20.7 | Montevideo | P.1/B.1.351* |
| CUY2-000266 | 18.3 | Montevideo | No-VOC |
| CUY2-000267 | 18.3 | Tacuarembó | P.1/B.1.351 |
| CUY2-000268 | 20.1 | Tacuarembó | P.1/B.1.351* |
| M525_UYRO | 19.0 | Rocha | P.1/B.1.351 |
| M530_UYRO | 14.6 | Rocha | ND |
| M531_UYRO | 12.3 | Rocha | ND |
| ND: No Data |  |  |  |

**Supplementary Table 2.**
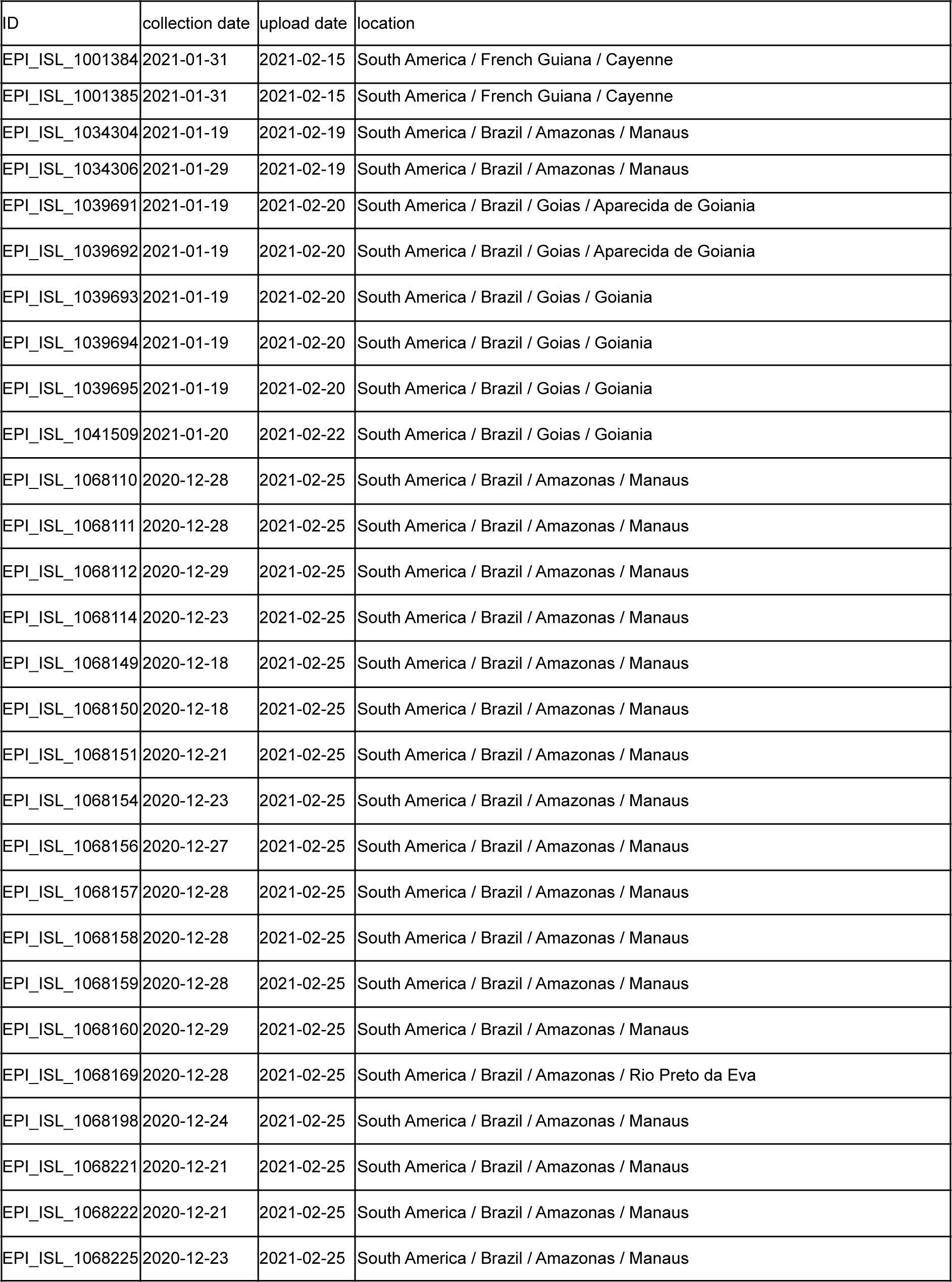
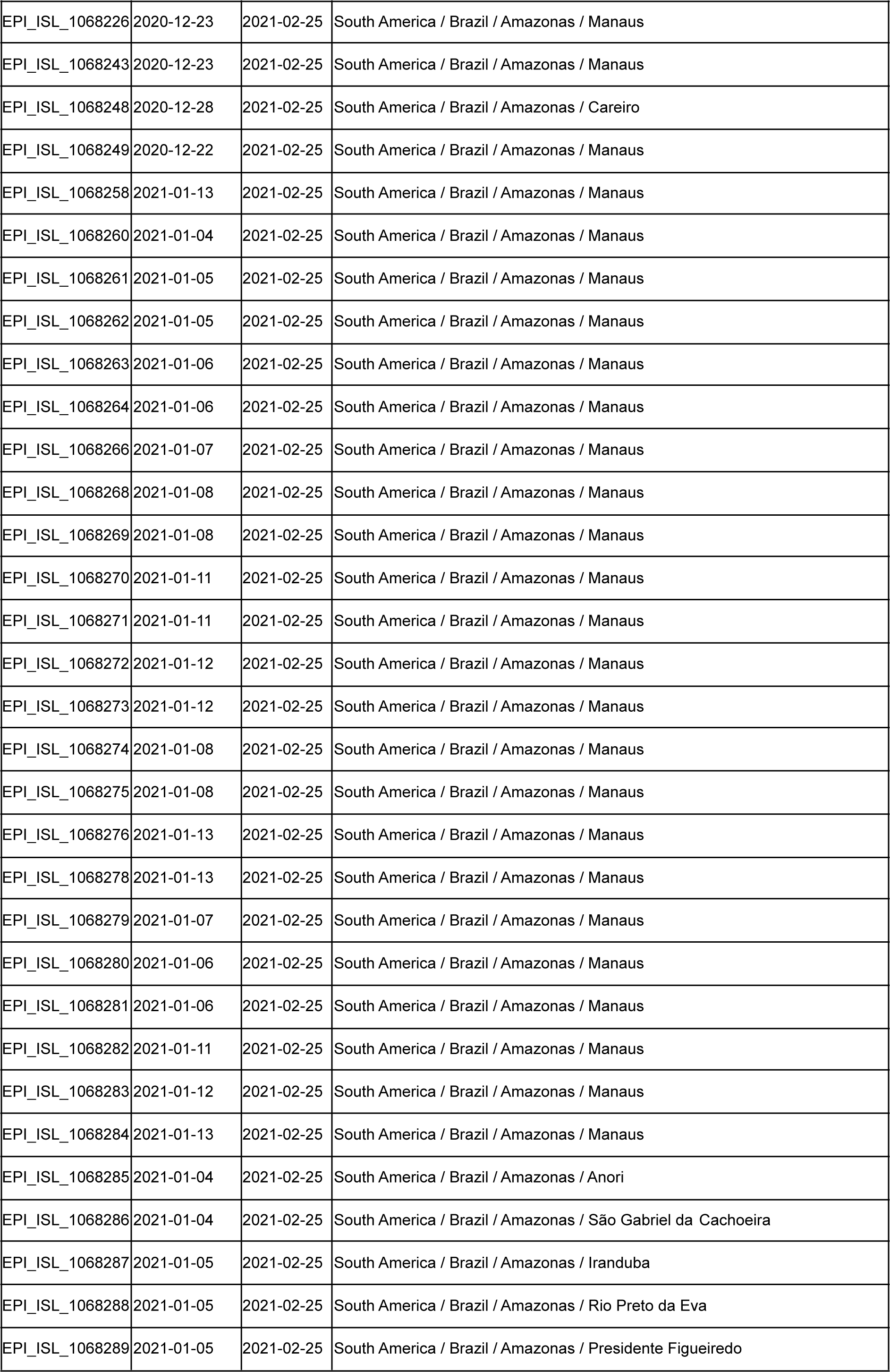
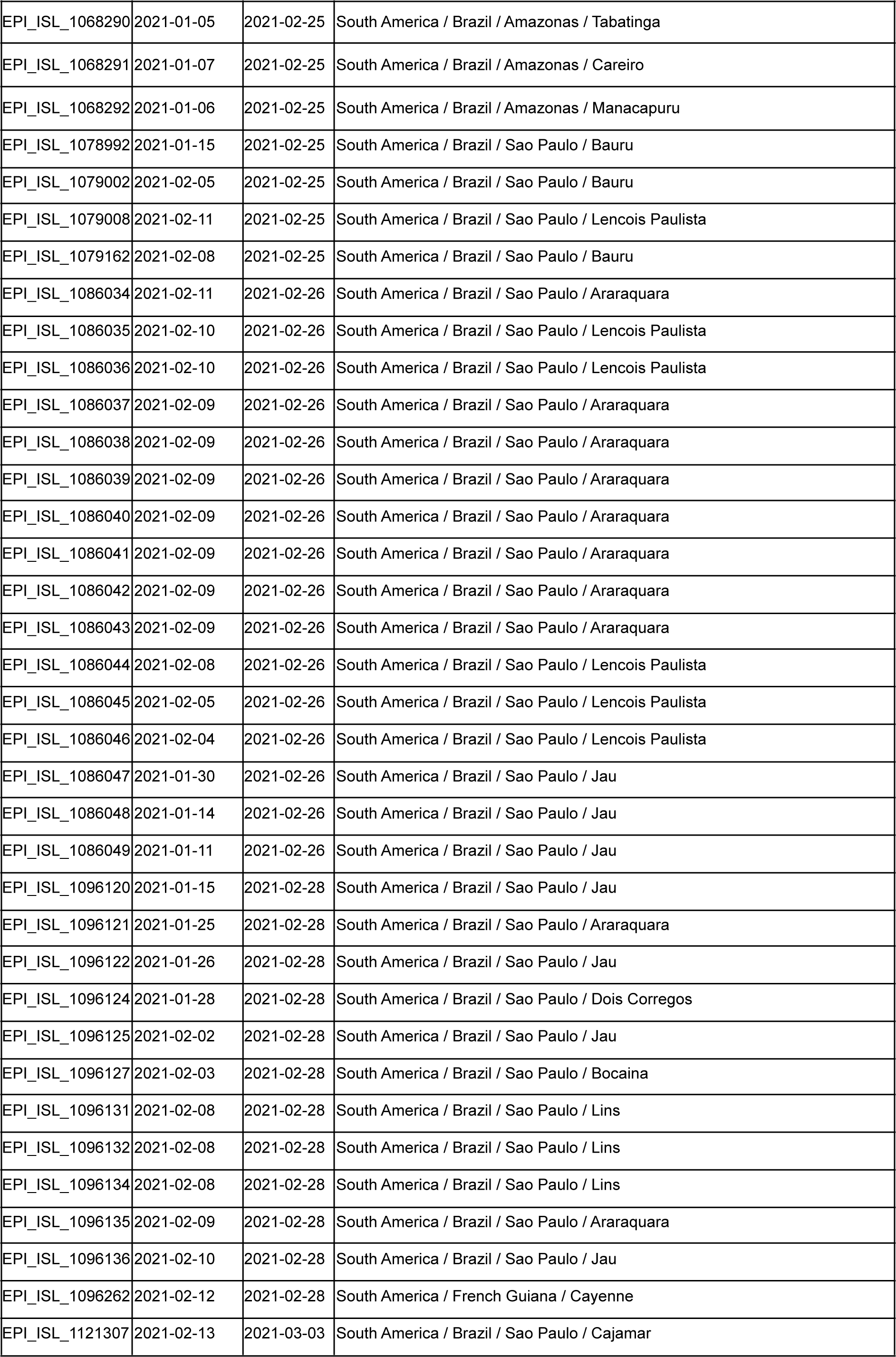
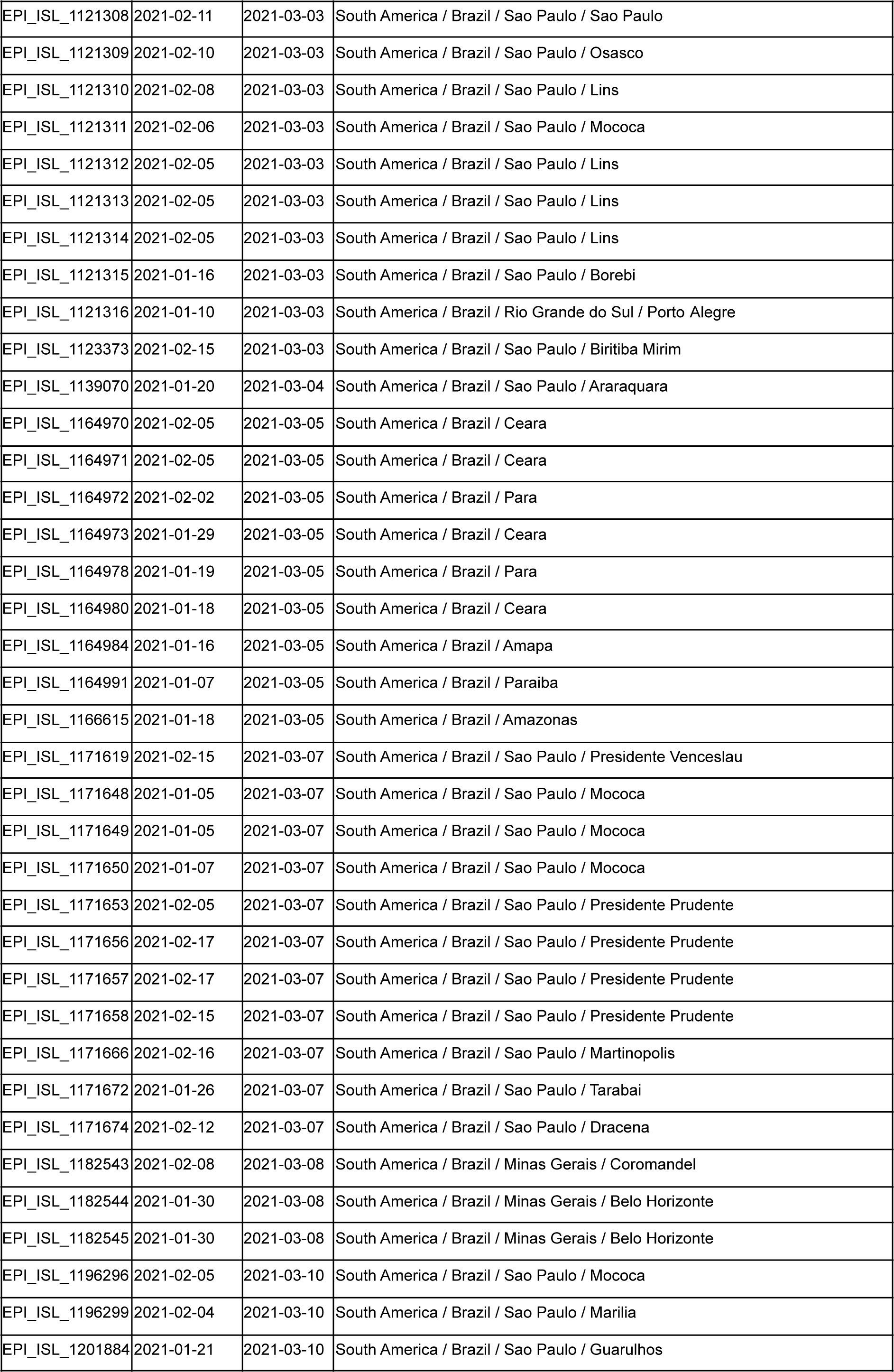
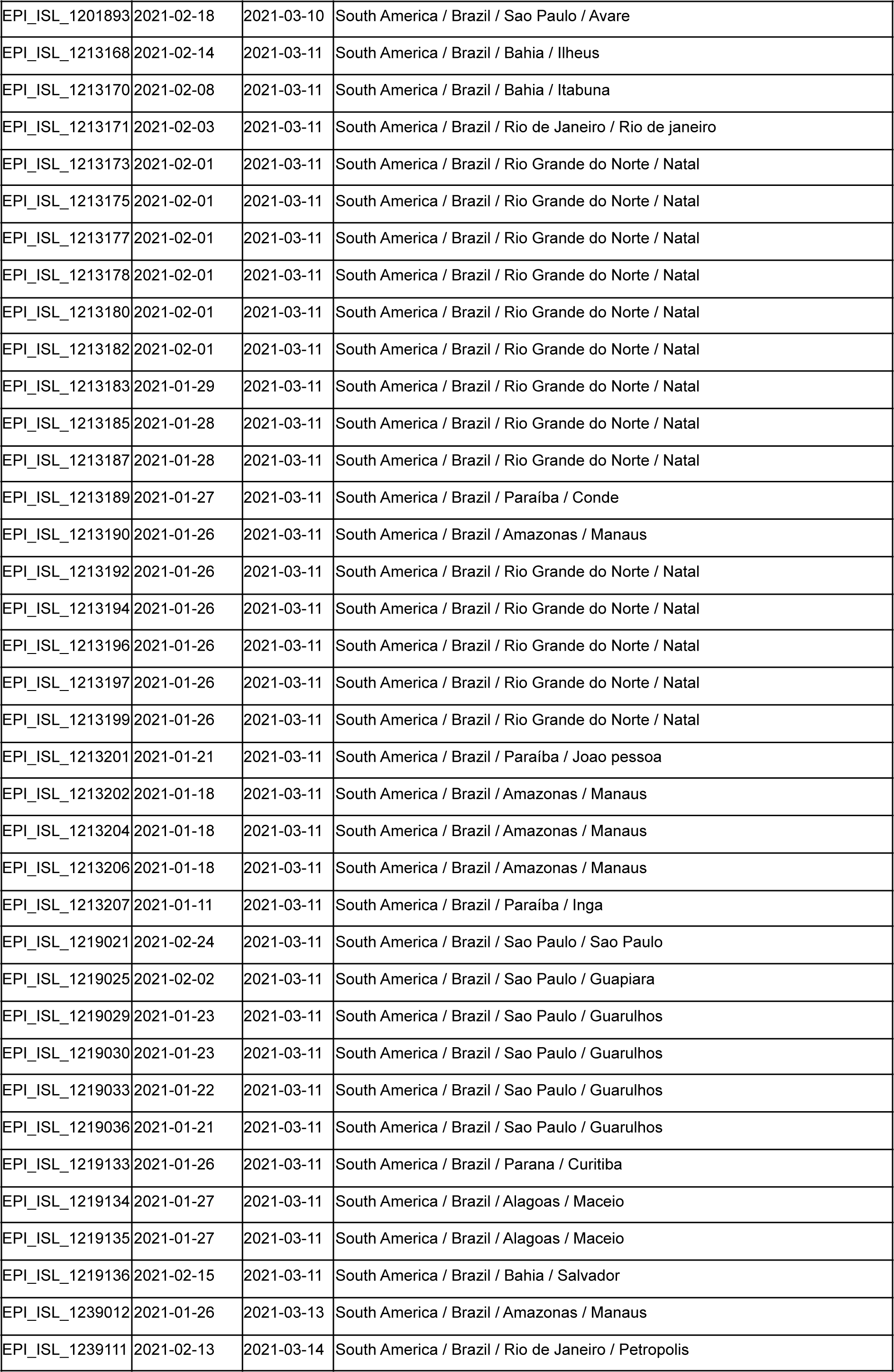
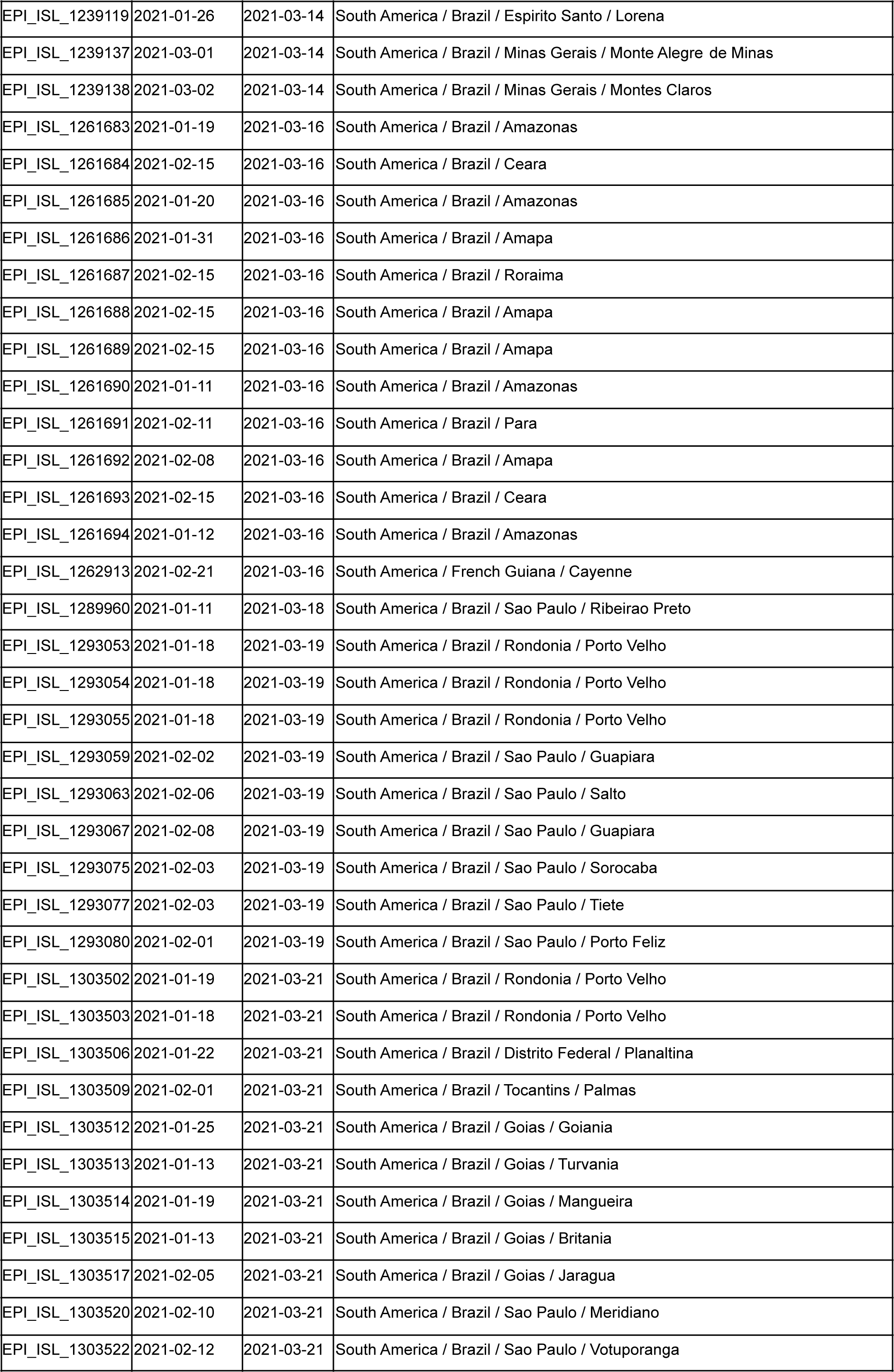
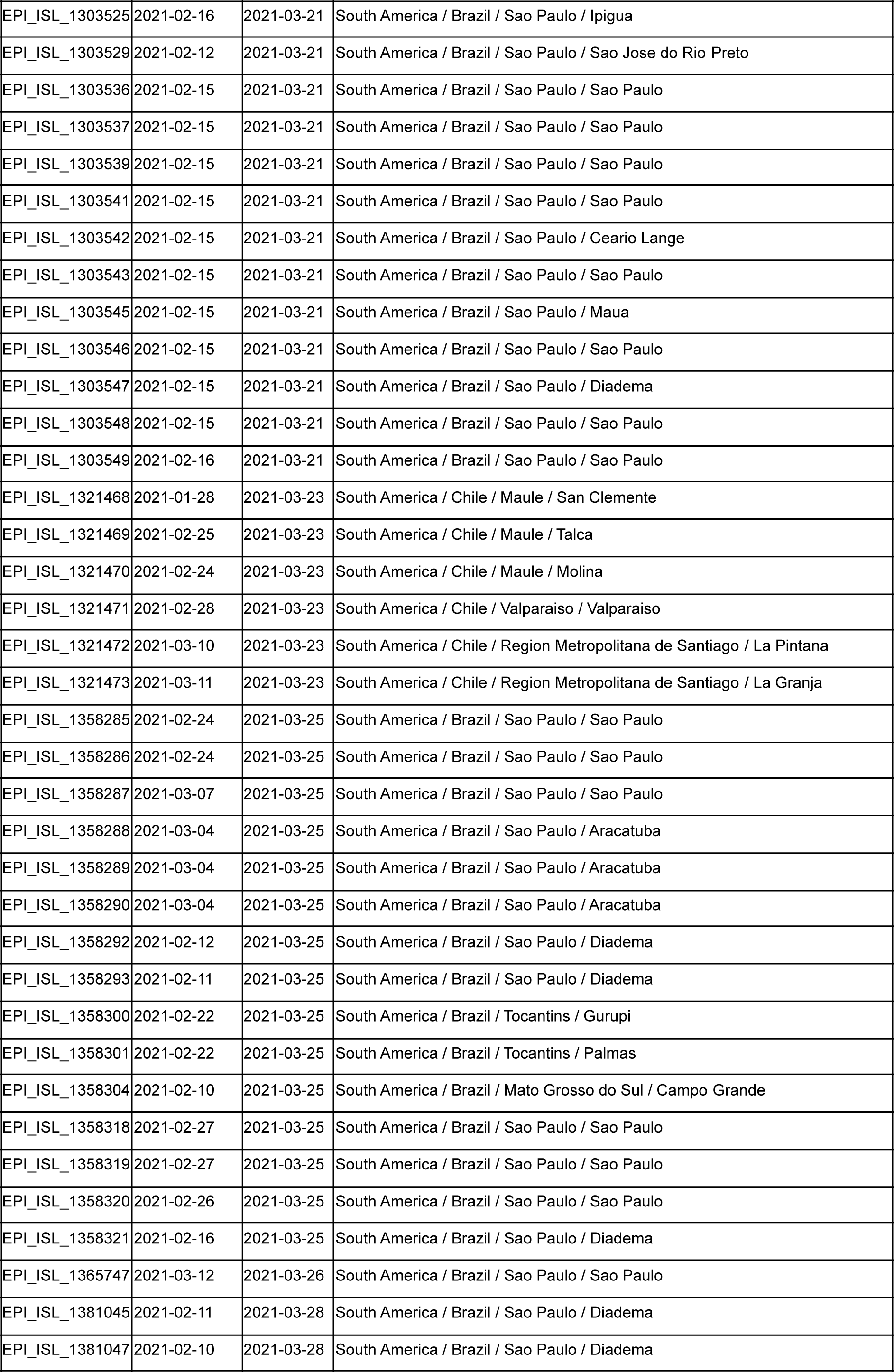
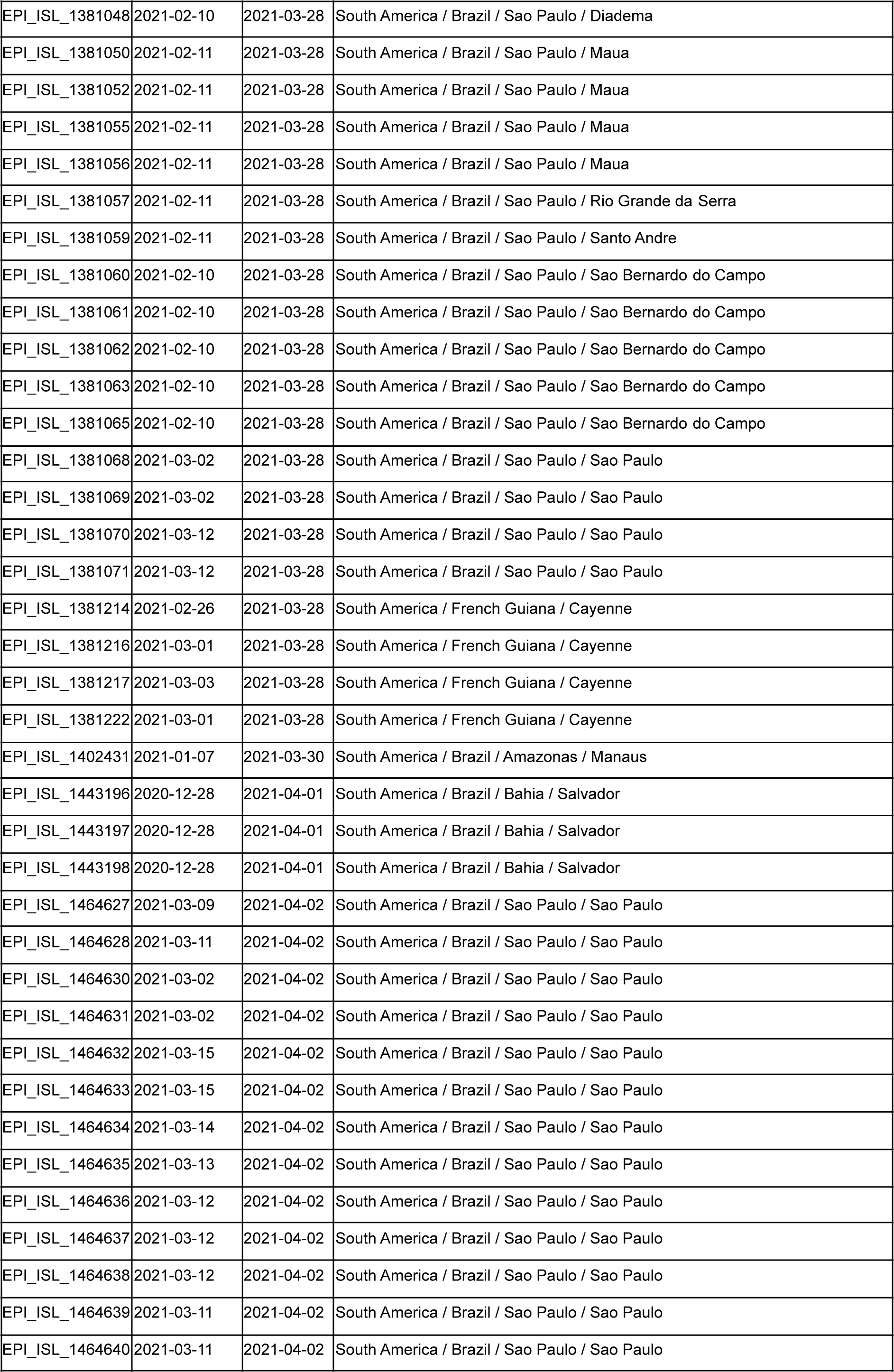
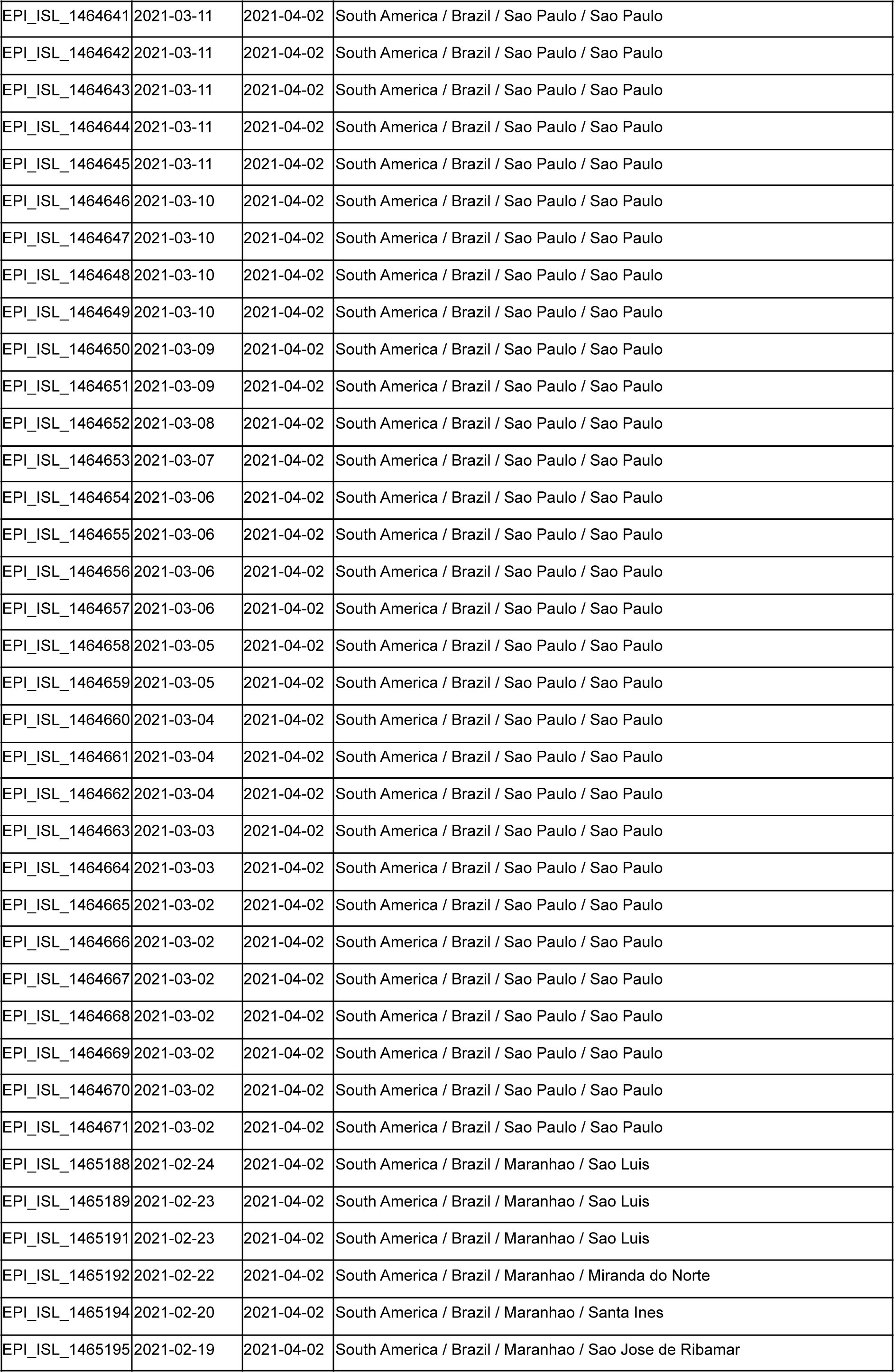
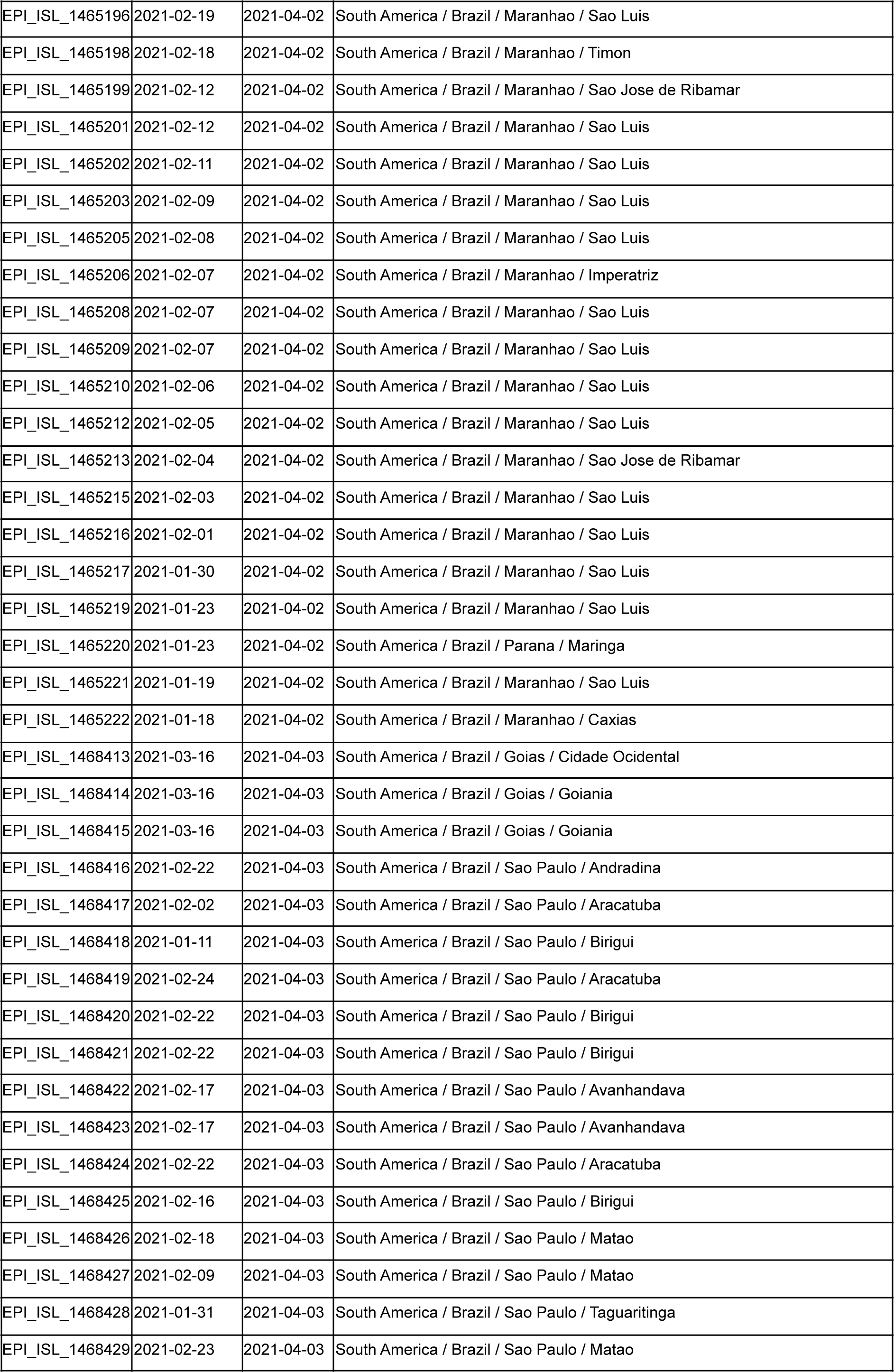
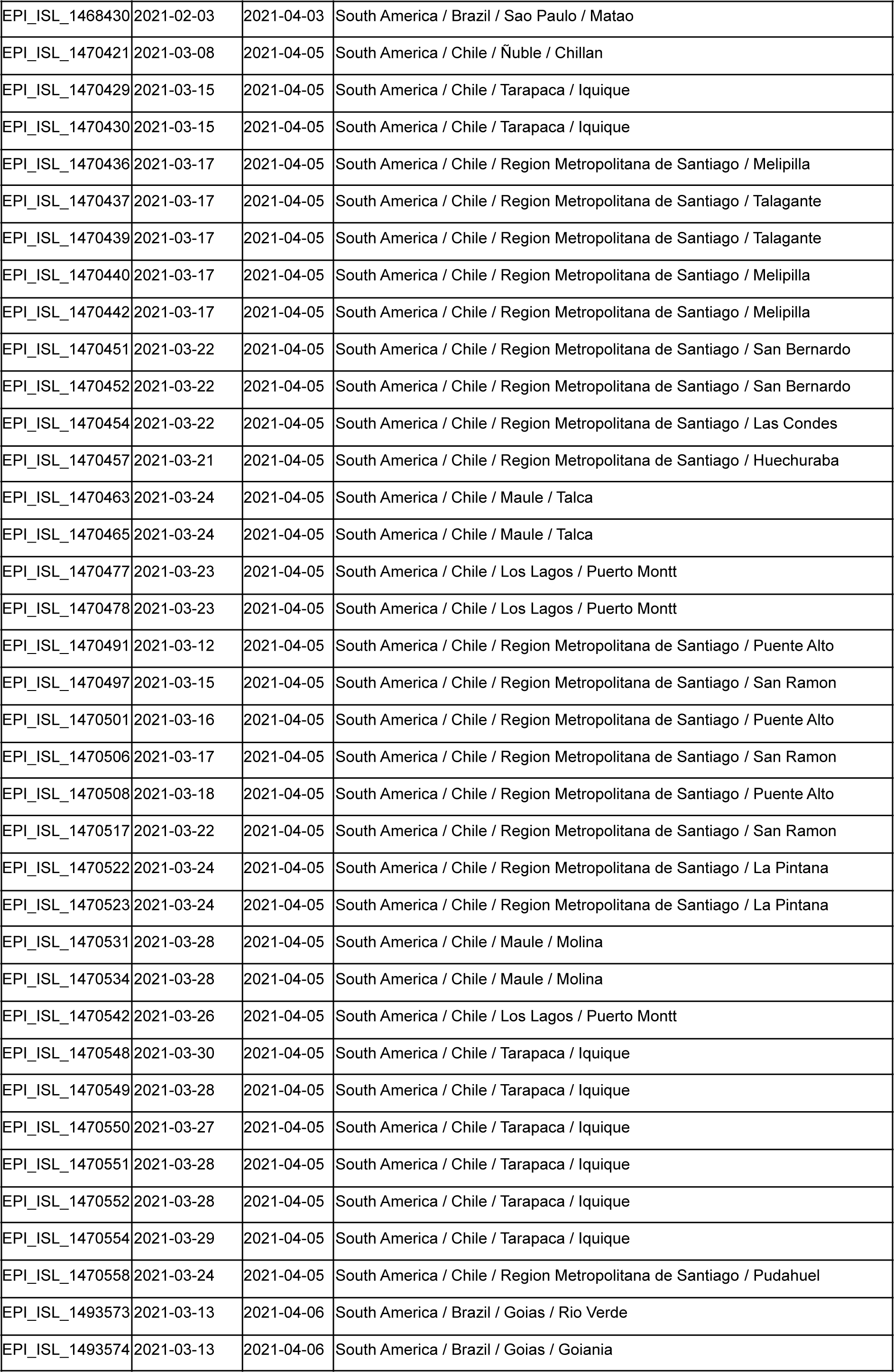
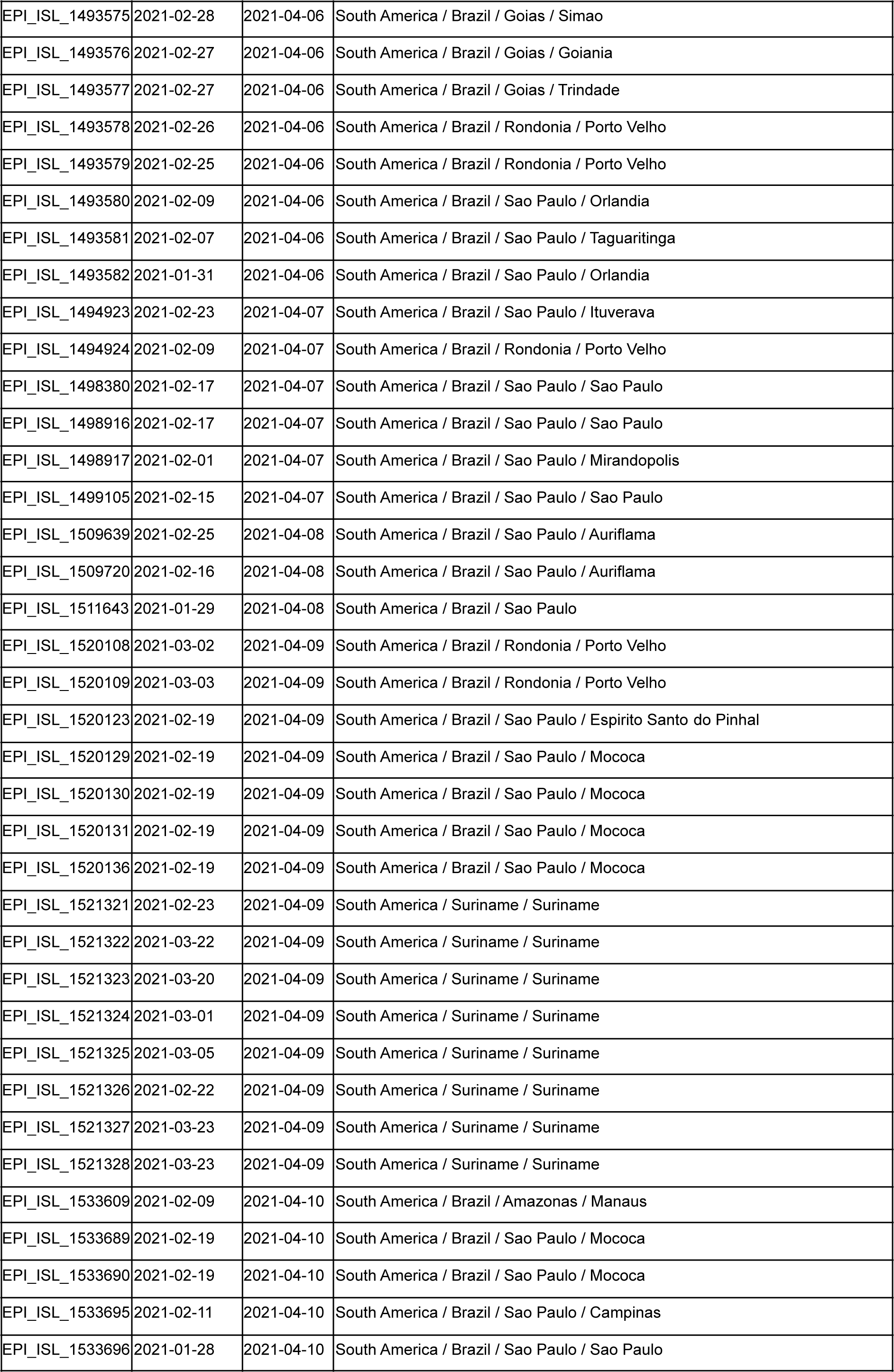
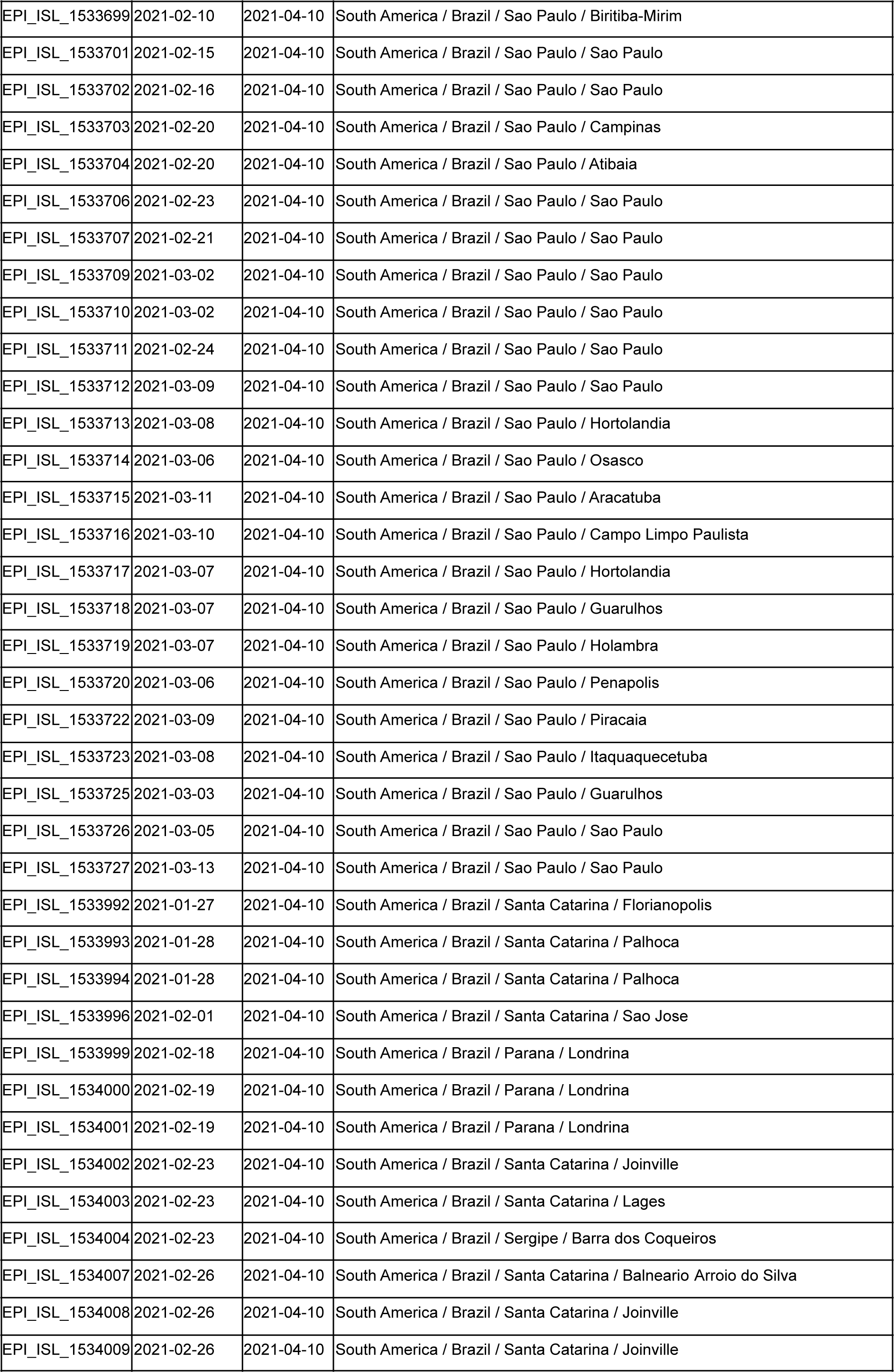
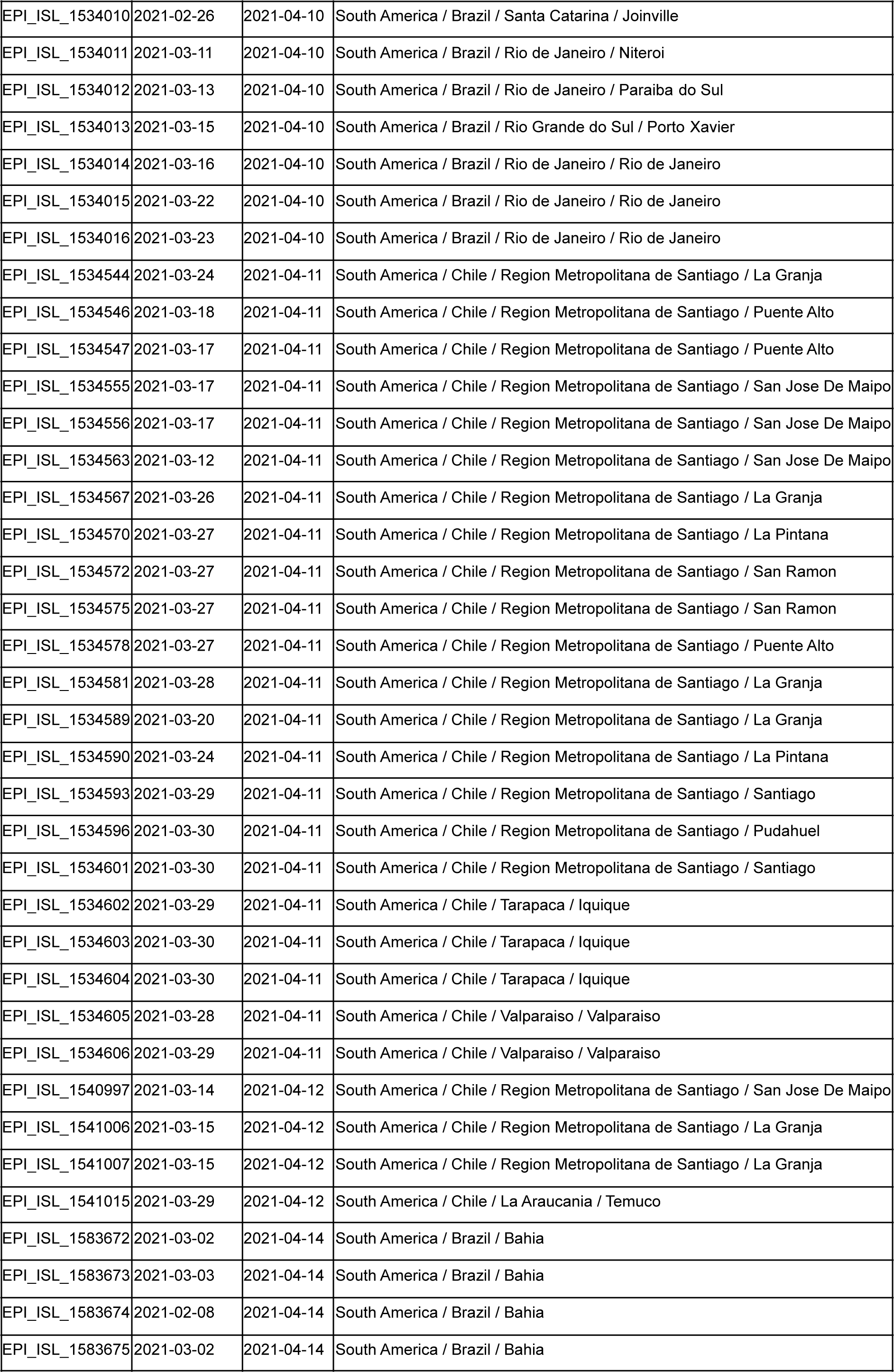
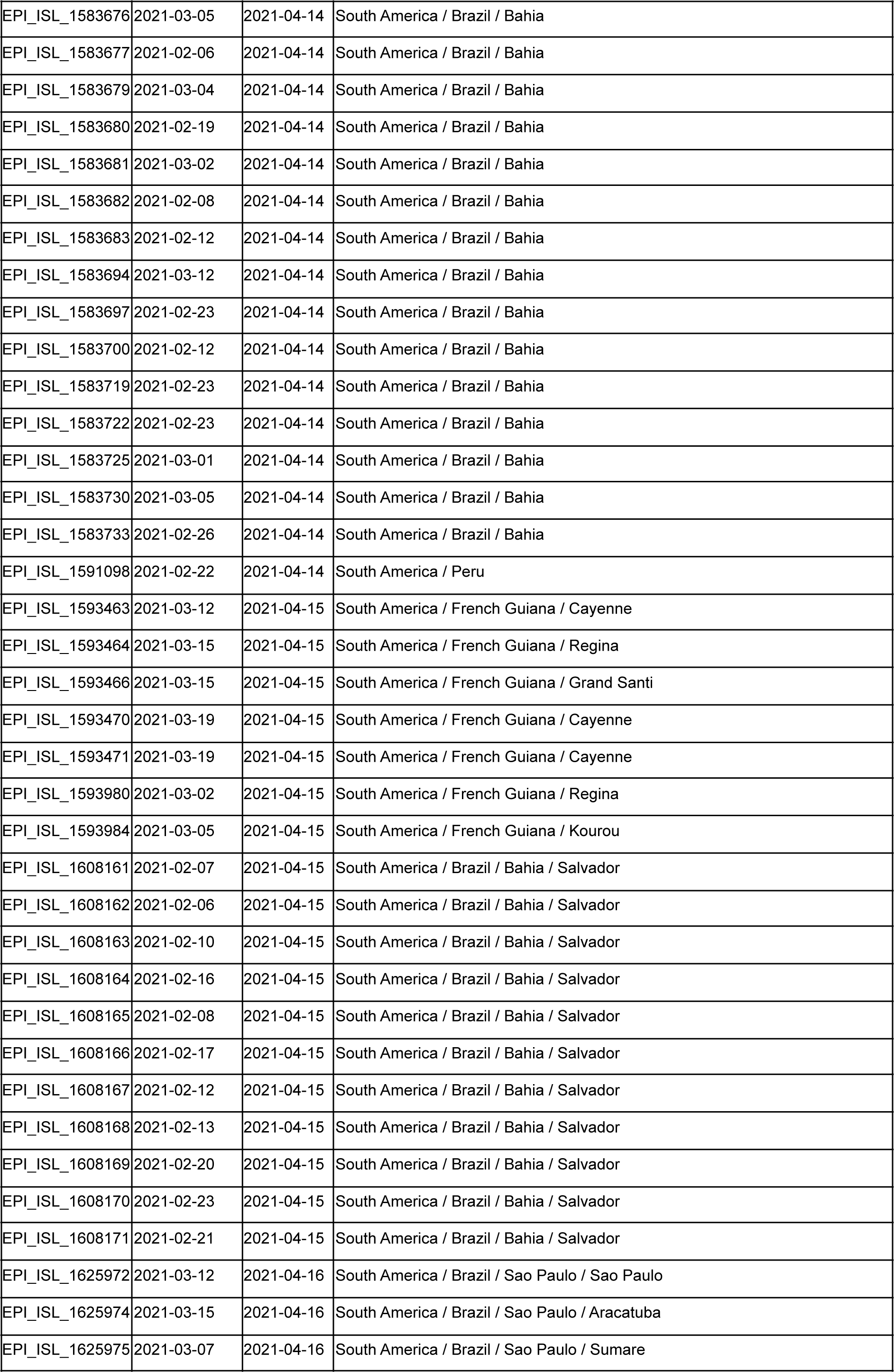
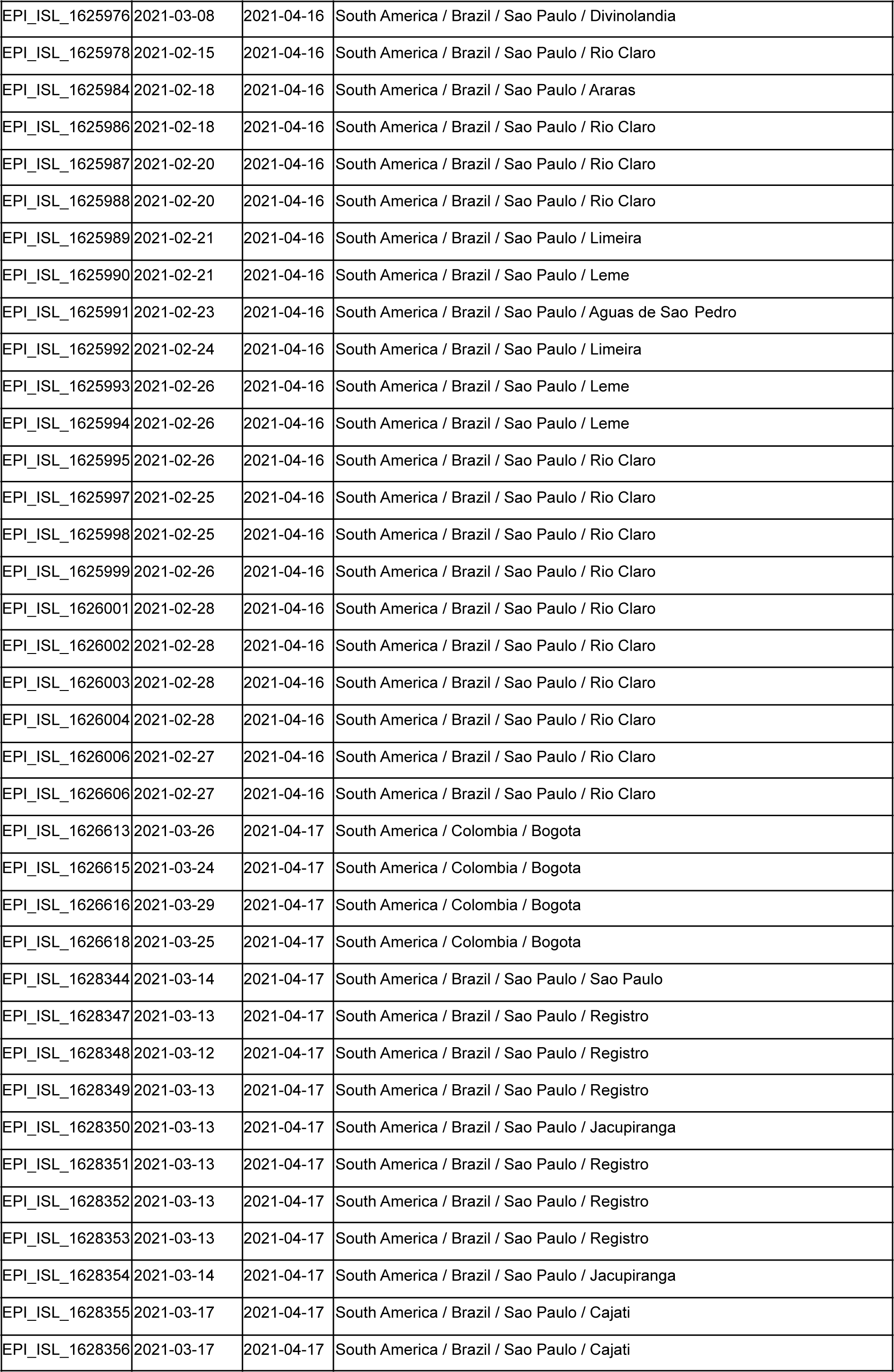
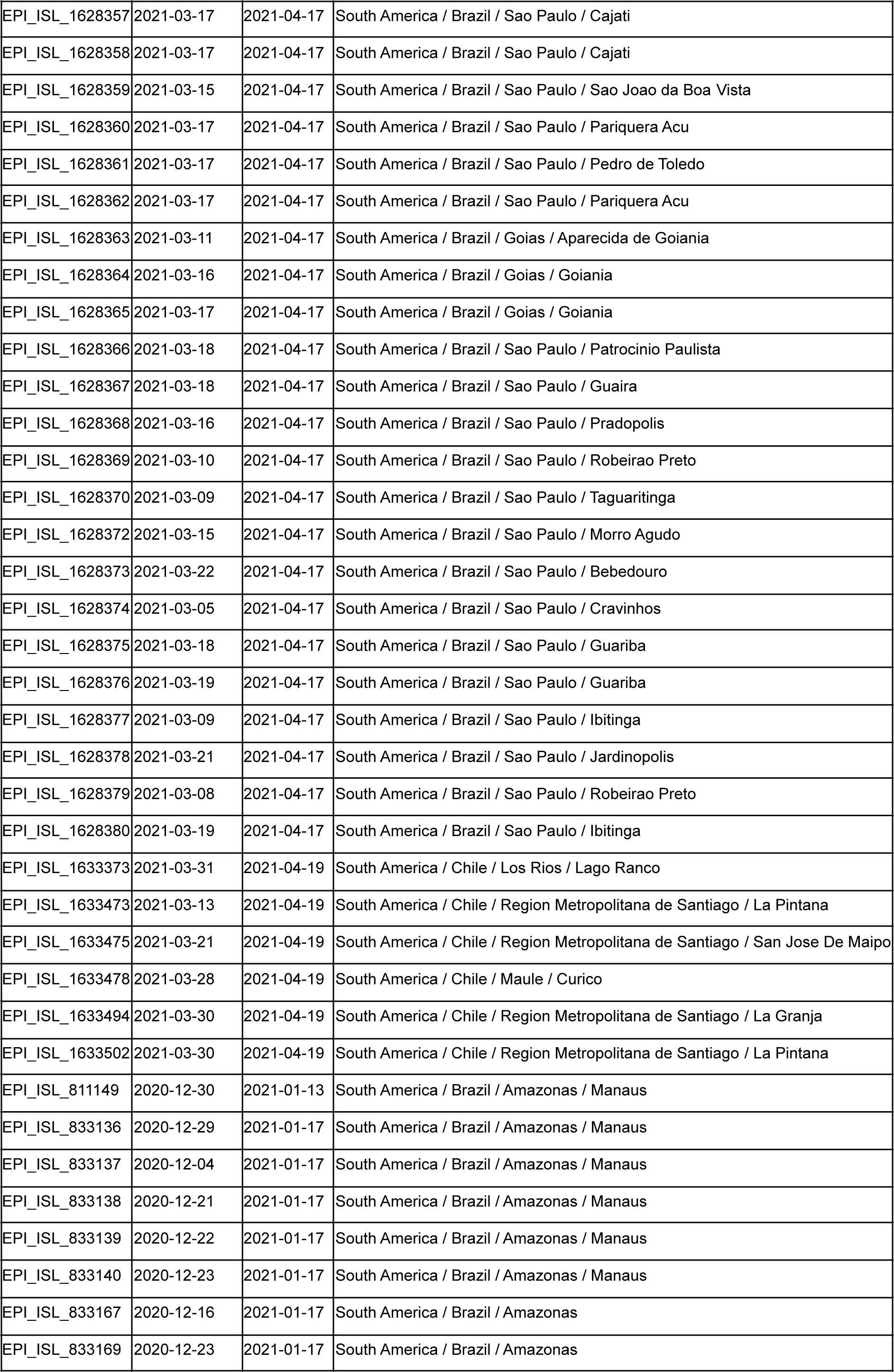
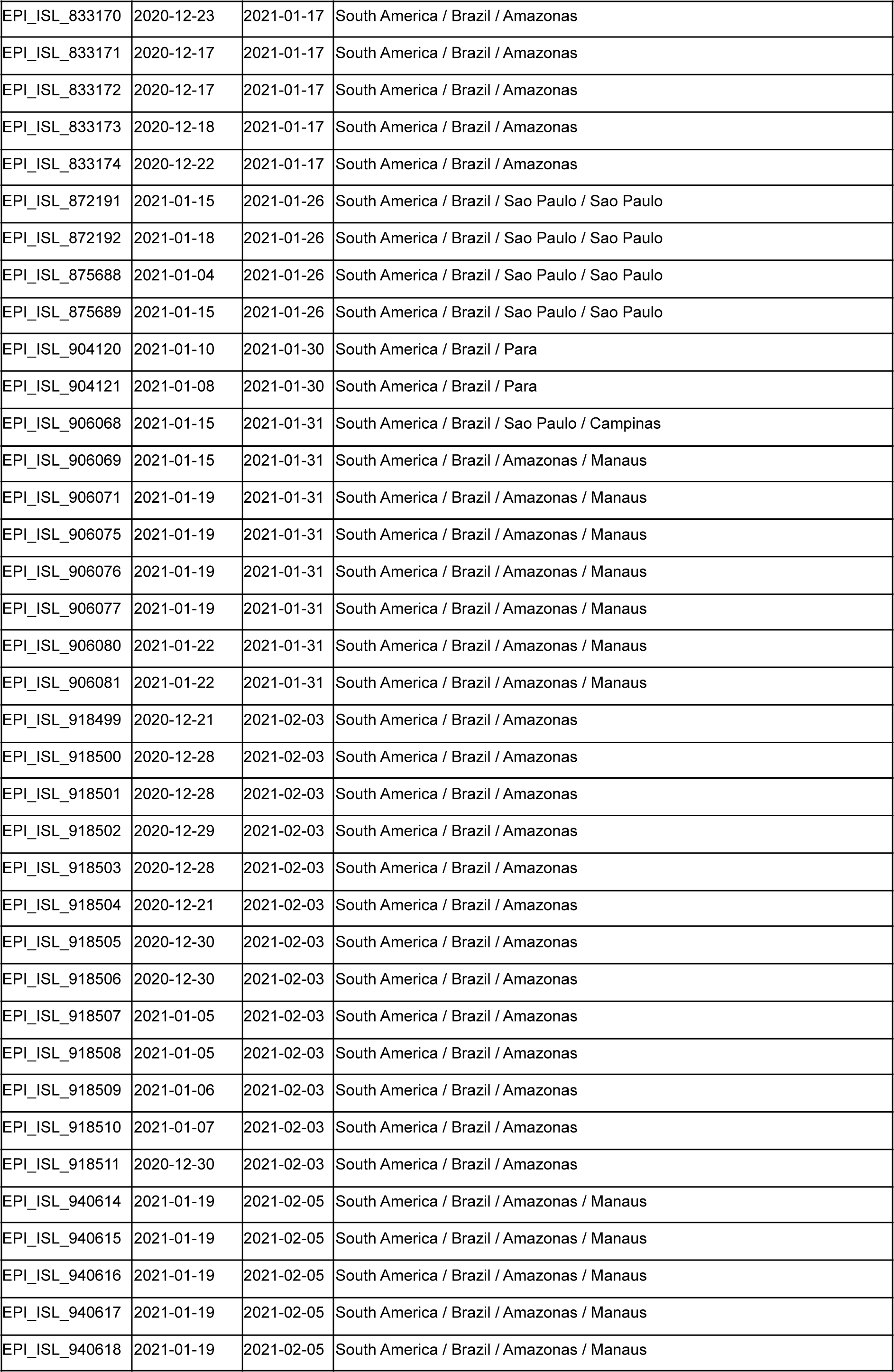
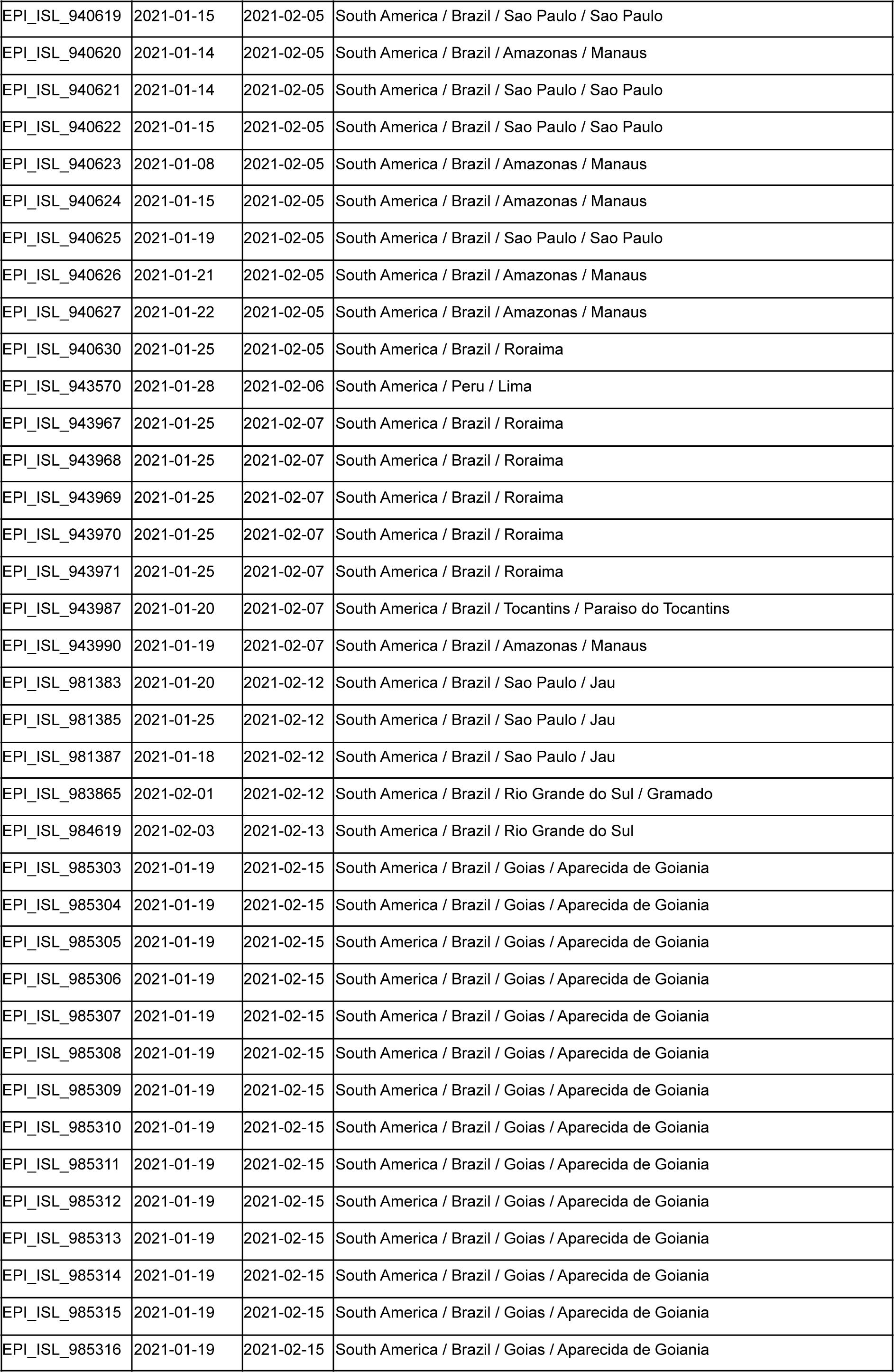
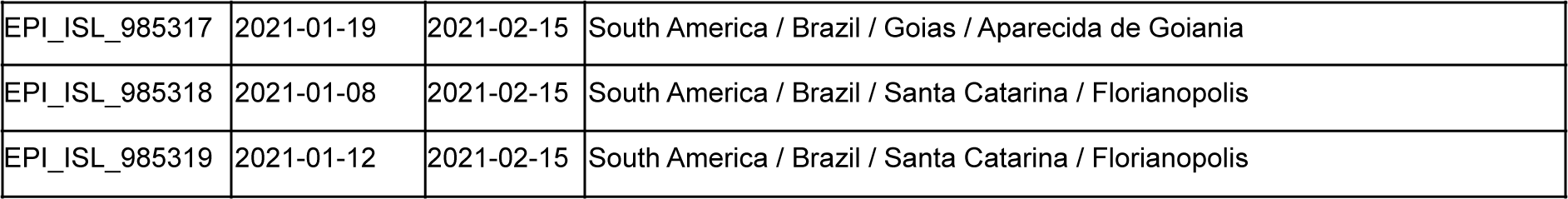
EpiCoV database identifiers (GISAID initiative) for SARS-CoV-2 downloaded sequences.

**Supplementary Table 3.**
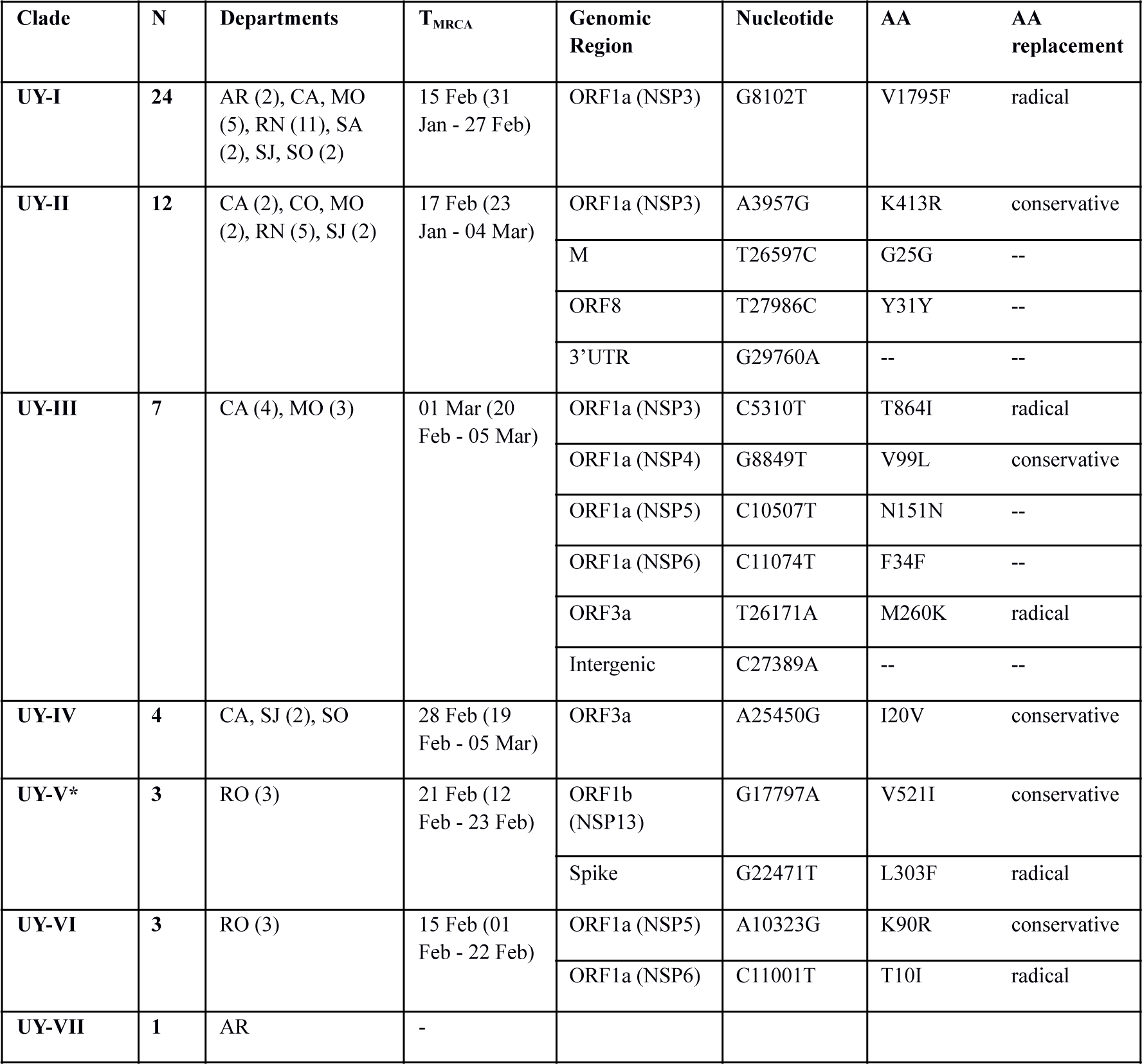
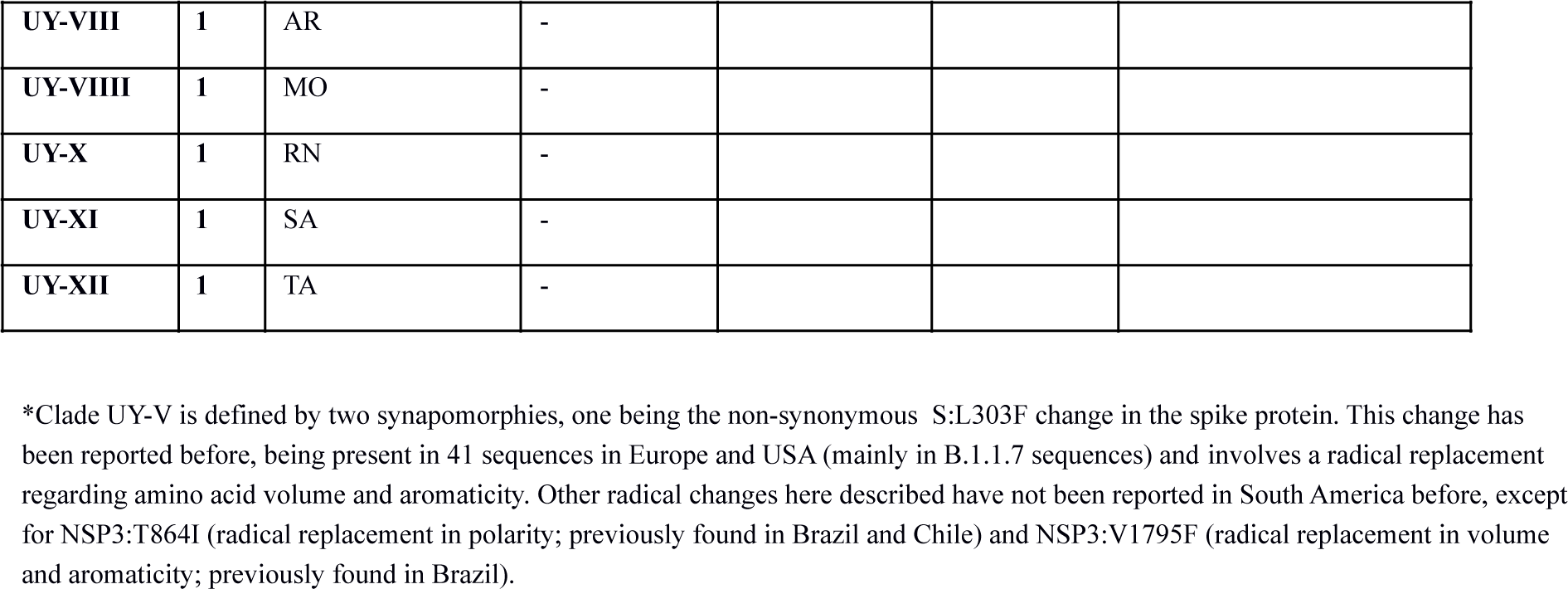
SARS-CoV-2 introductions of P.1 lineage in Uruguay and synapomorphic sites defining each Uruguayan clade. Columns show the identified Uruguayan P.1 clade, the number of samples per clade, the Uruguayan departements where de clade is present (and number of samples), the T_MRCA_ as estimated by BEAST (and range of the 95% confidence interval), and a description of the synapomorphies defining each clade (for those with at least three samples).

**Supplementary Table 4.**
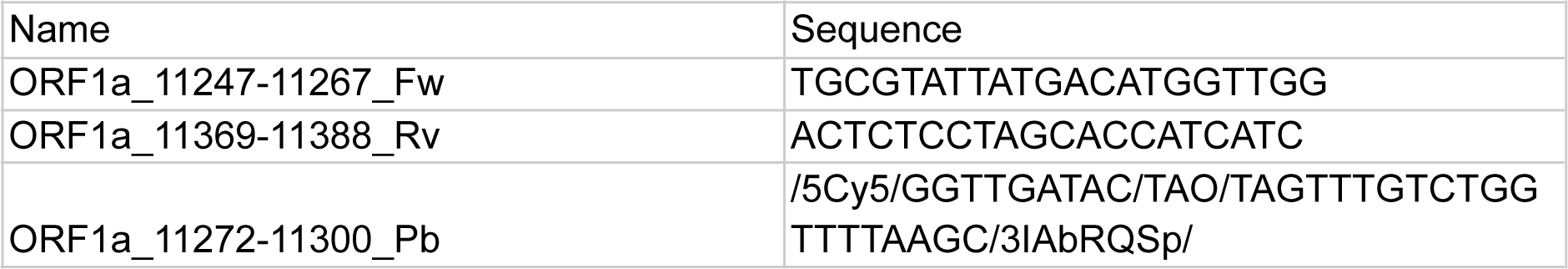
Primers and probe designed for ORF1ab drop-out qPCR.

## 4. Supplementary Figures

**Supplementary Figure 1:**
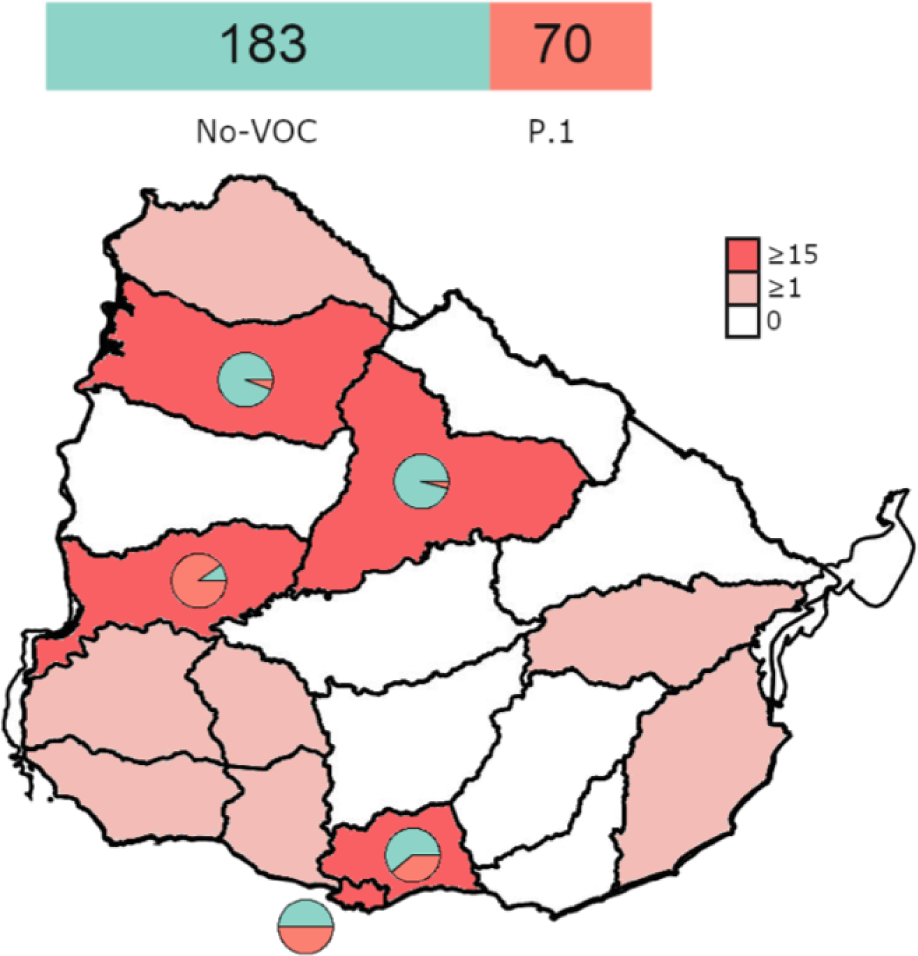
Barplot showing proportions of P.1 and no-VOCs. Total numbers of samples in each category are shown (top). Uruguayan map with P.1 locations marked in red. Locations with more than 15 samples are marked in dark red and between one and 14 with light red. Pie charts indicate proportions of VOC and No-VOCs in departments with a high number of samples (more than 15): VOC in red and no-VOCs in green.

**Supplementary Figure 2.**
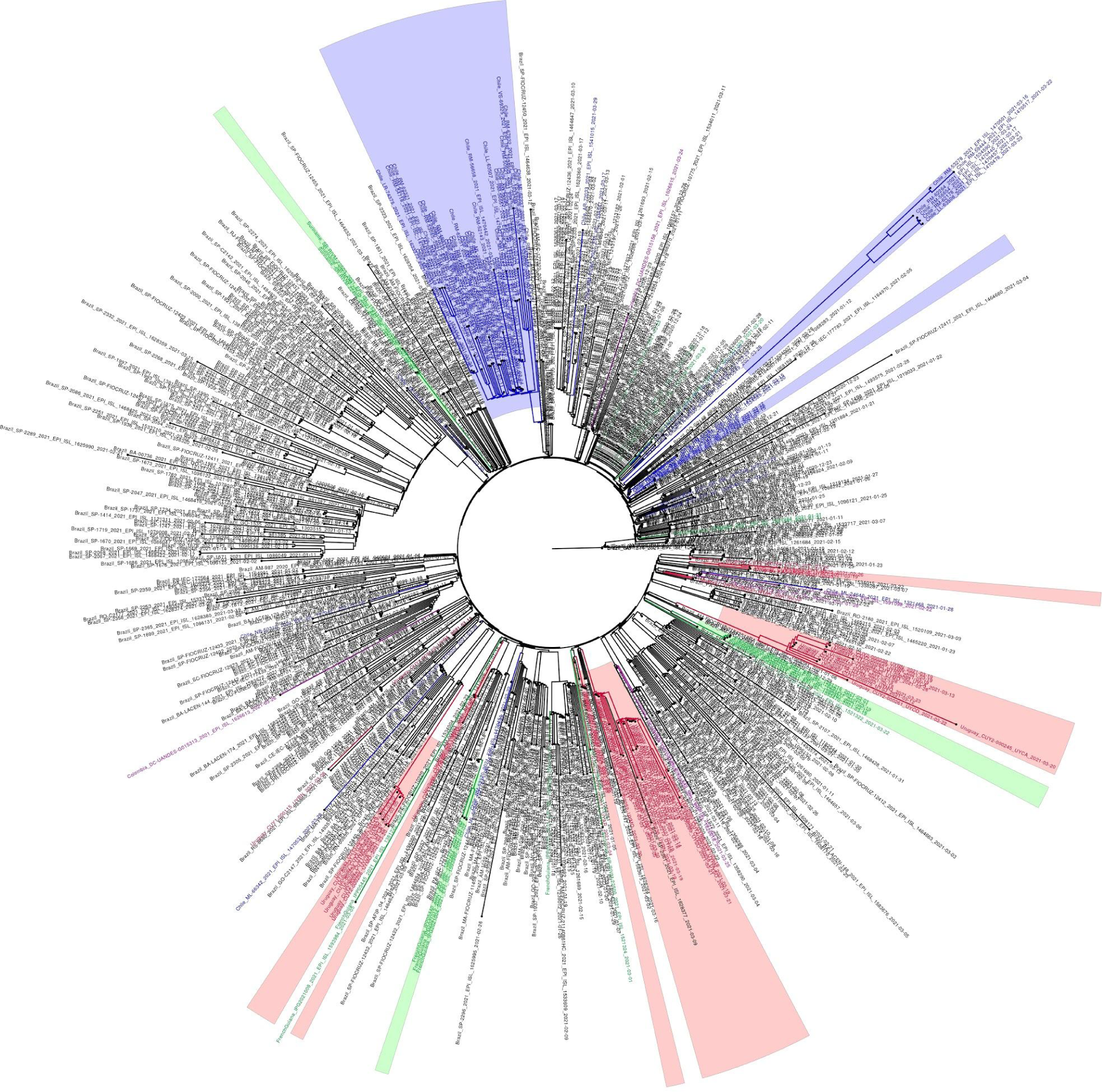
Maximum-likelihood tree of 59 Uruguayan and 691 South American P.1 whole genome sequences. The tree was rooted with the oldest P.1 sequence (EPI_ISL_833137). Brazilian sequences are kept in black. Uruguayan sequences are shown in red. While six sequences are singletons, 53 sequences gather in six clades of size three to 24 (red areas). Chilean sequences and clades are shown in blue, French Guiana and Suriname sequences and clades in green. Sequences from Colombia and Peru are colored violet.

**Supplementary Figure 3.**
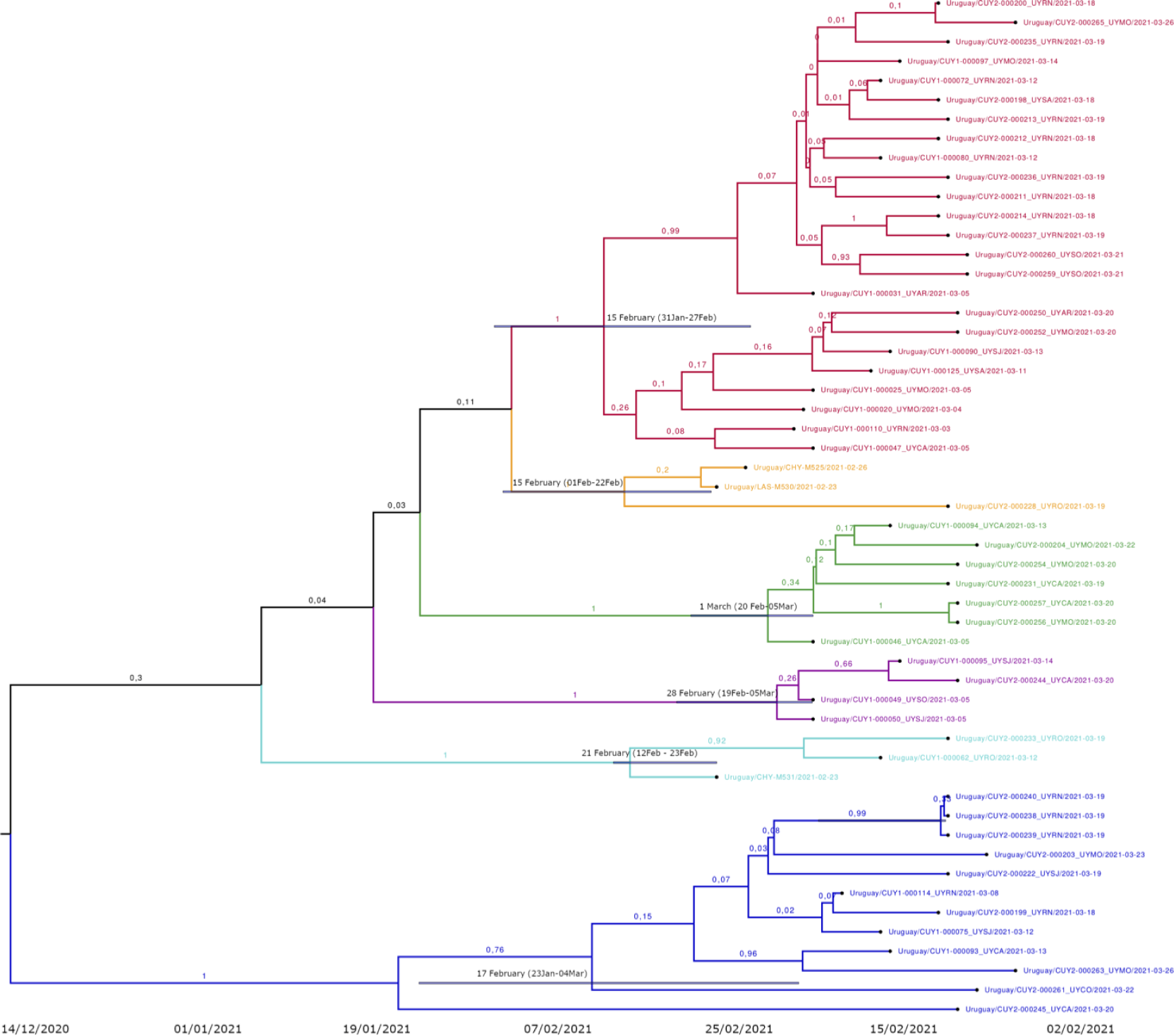
Temporal dissemination of SARS-CoV-2 Uruguayan clades. Six Uruguayan clades inferred on the time-scaled bayesian phylogeographic MCC tree. T_MRCA_ with 95% confidence interval at ancestral nodes are shown. Clades are colored: UY-I: red, UY-II: blue, UY-III: green, UY-IV: violet, UY-V: aquamarine, UY-VI: yellow.

## 5. Supplementary GISAID Acknowledgment Table

We gratefully acknowledge the following Authors from the Originating laboratories responsible for obtaining the specimens, as well as the Submitting laboratories where the genome data were generated and shared via GISAID, on which this research is based.

All Submitters of data may be contacted directly via www.gisaid.org

Authors are sorted alphabetically.

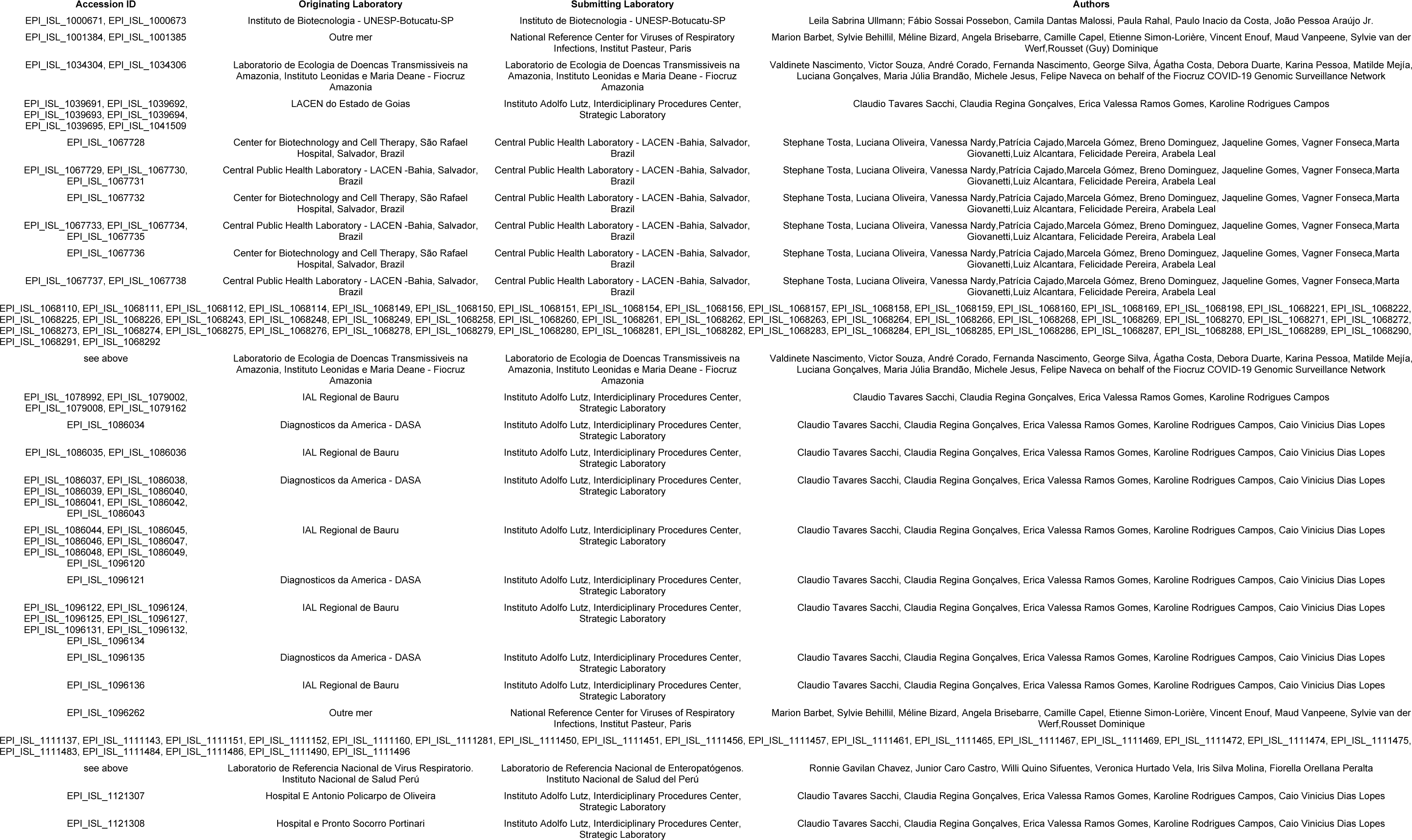
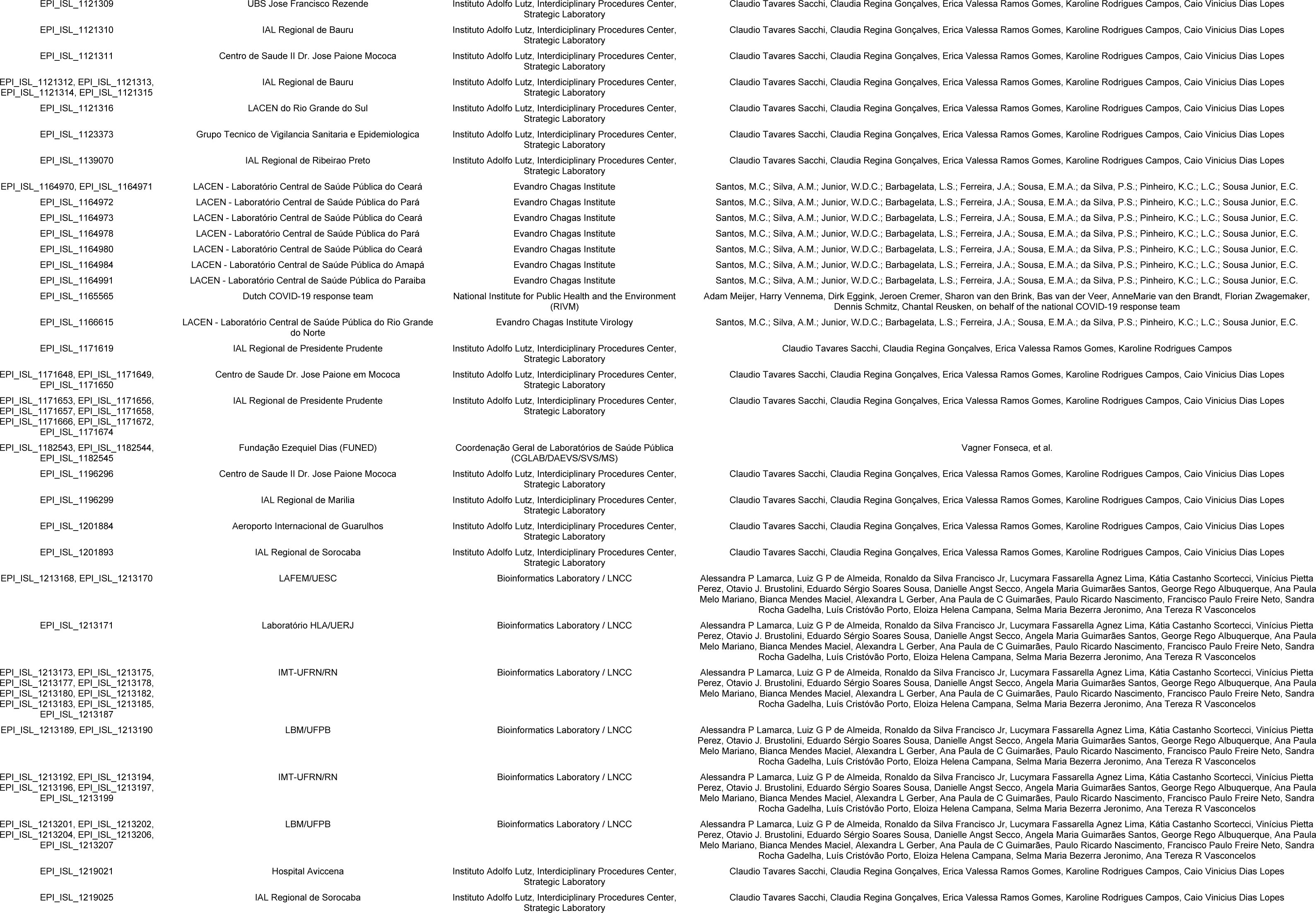
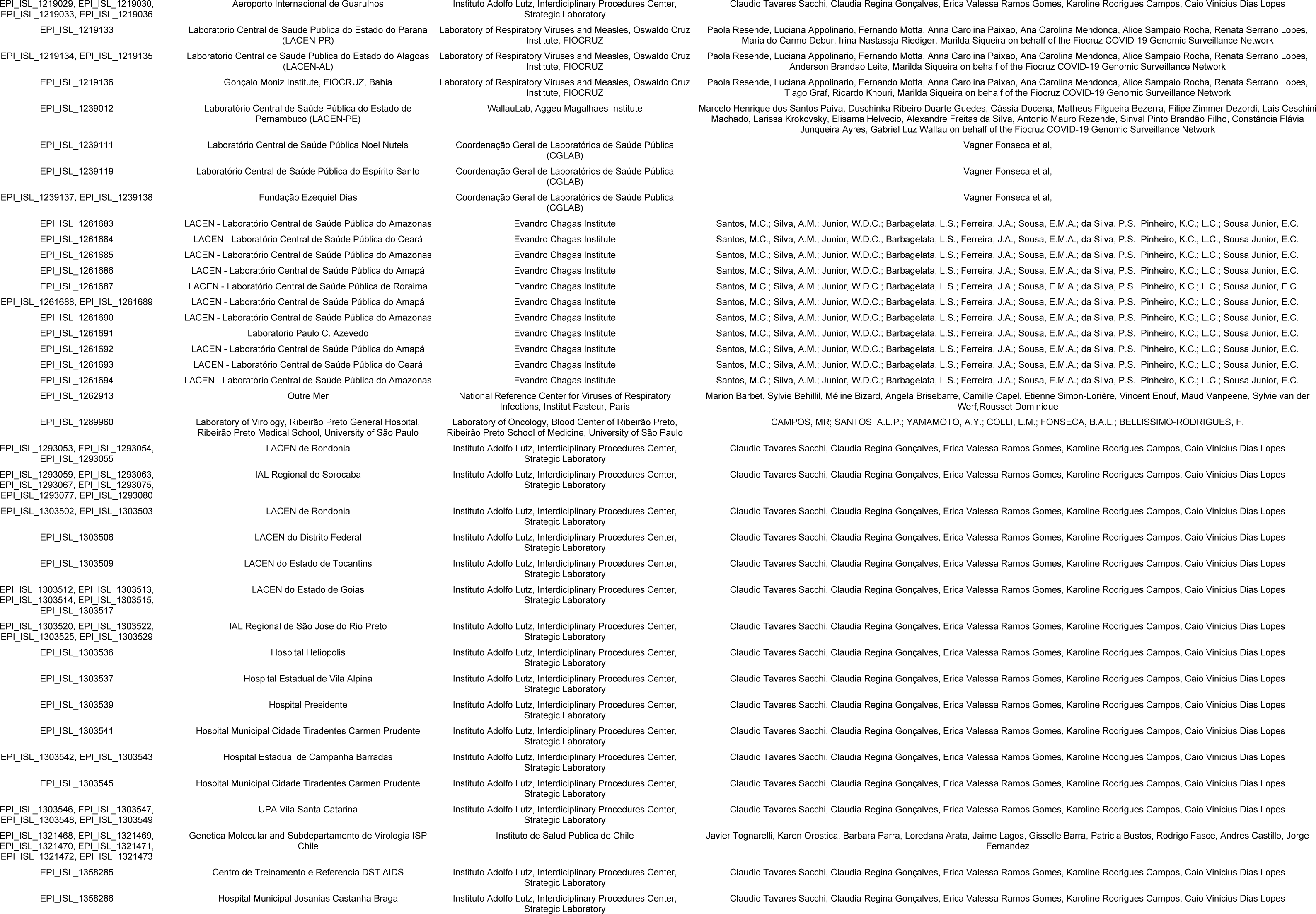
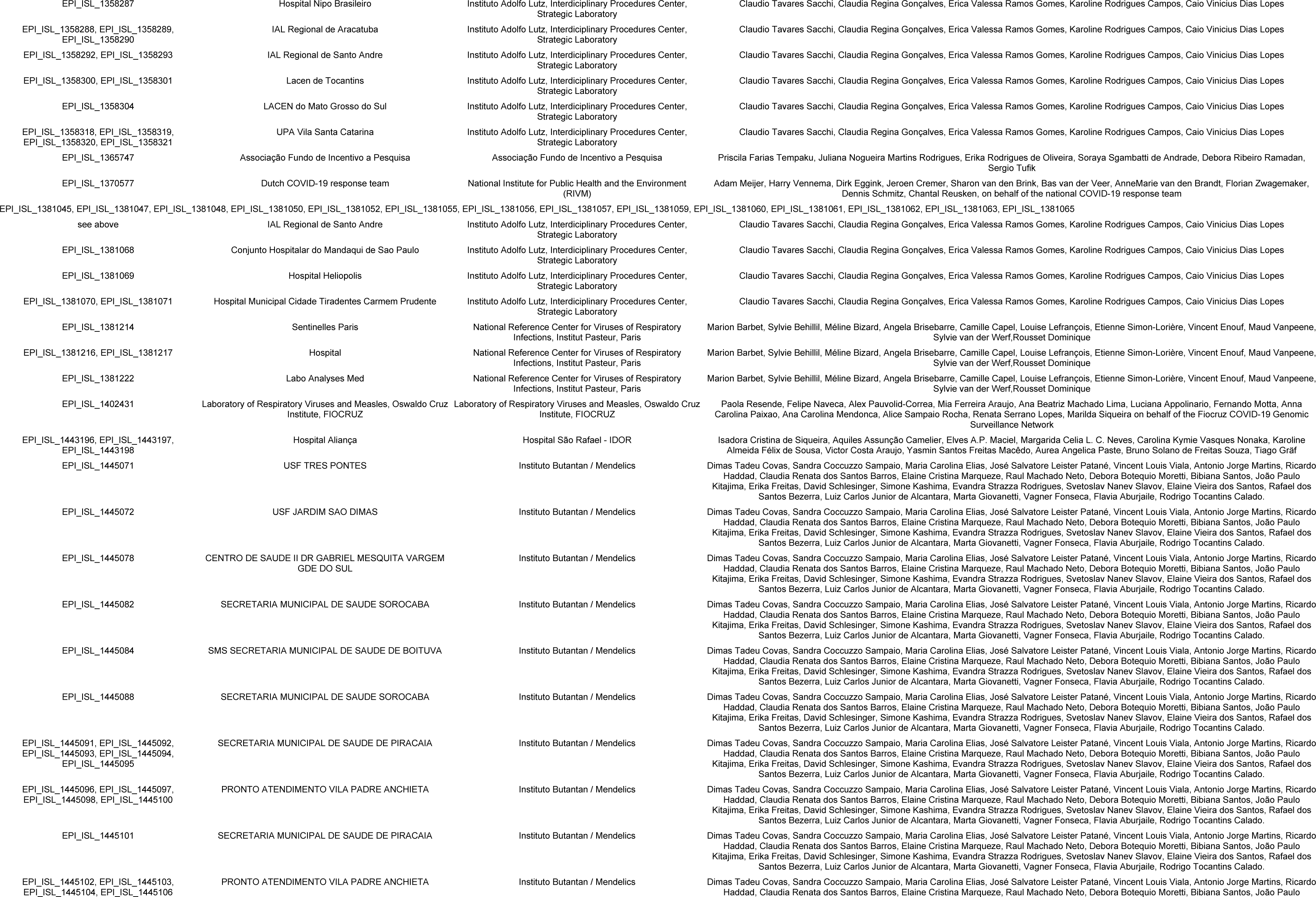
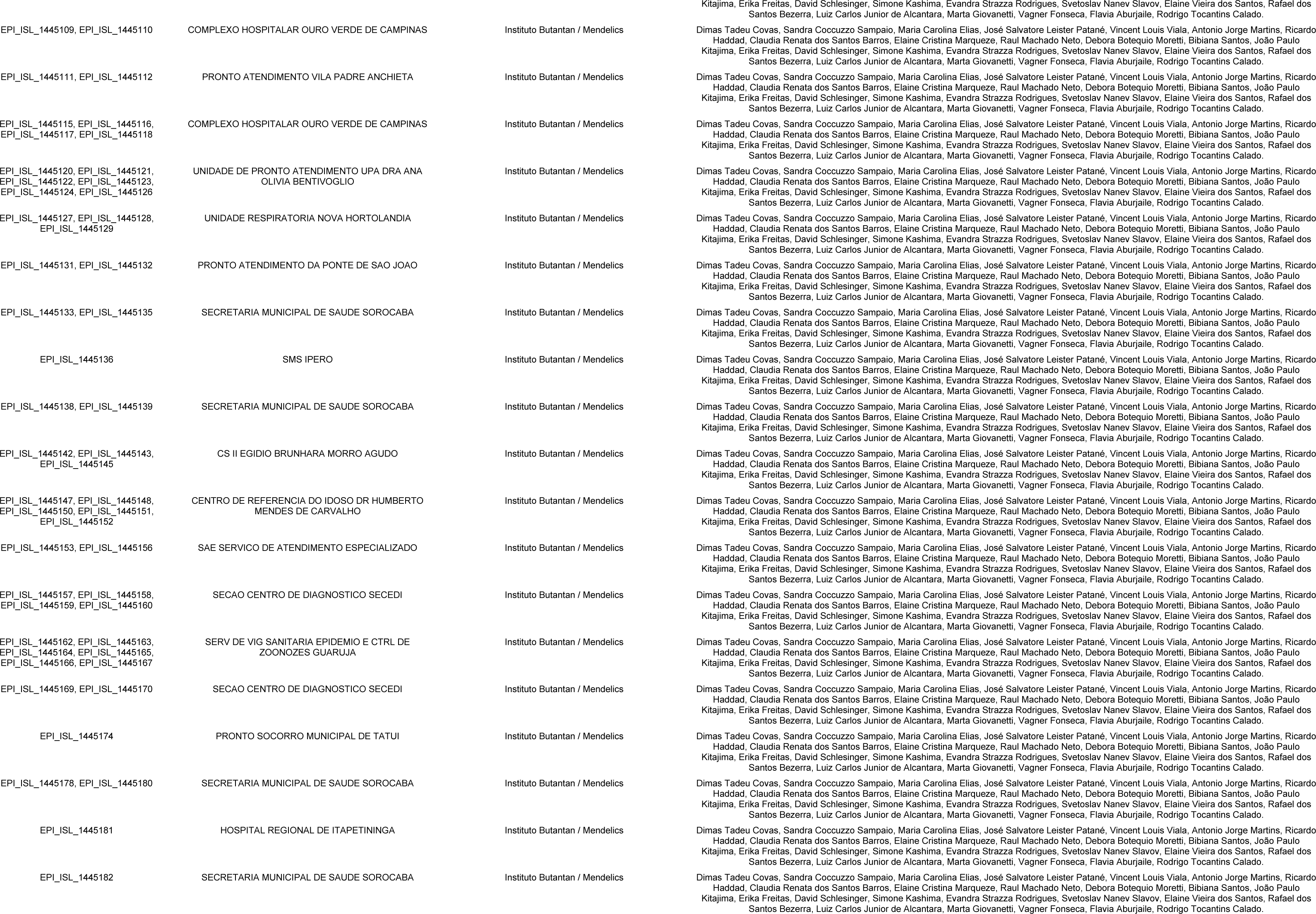
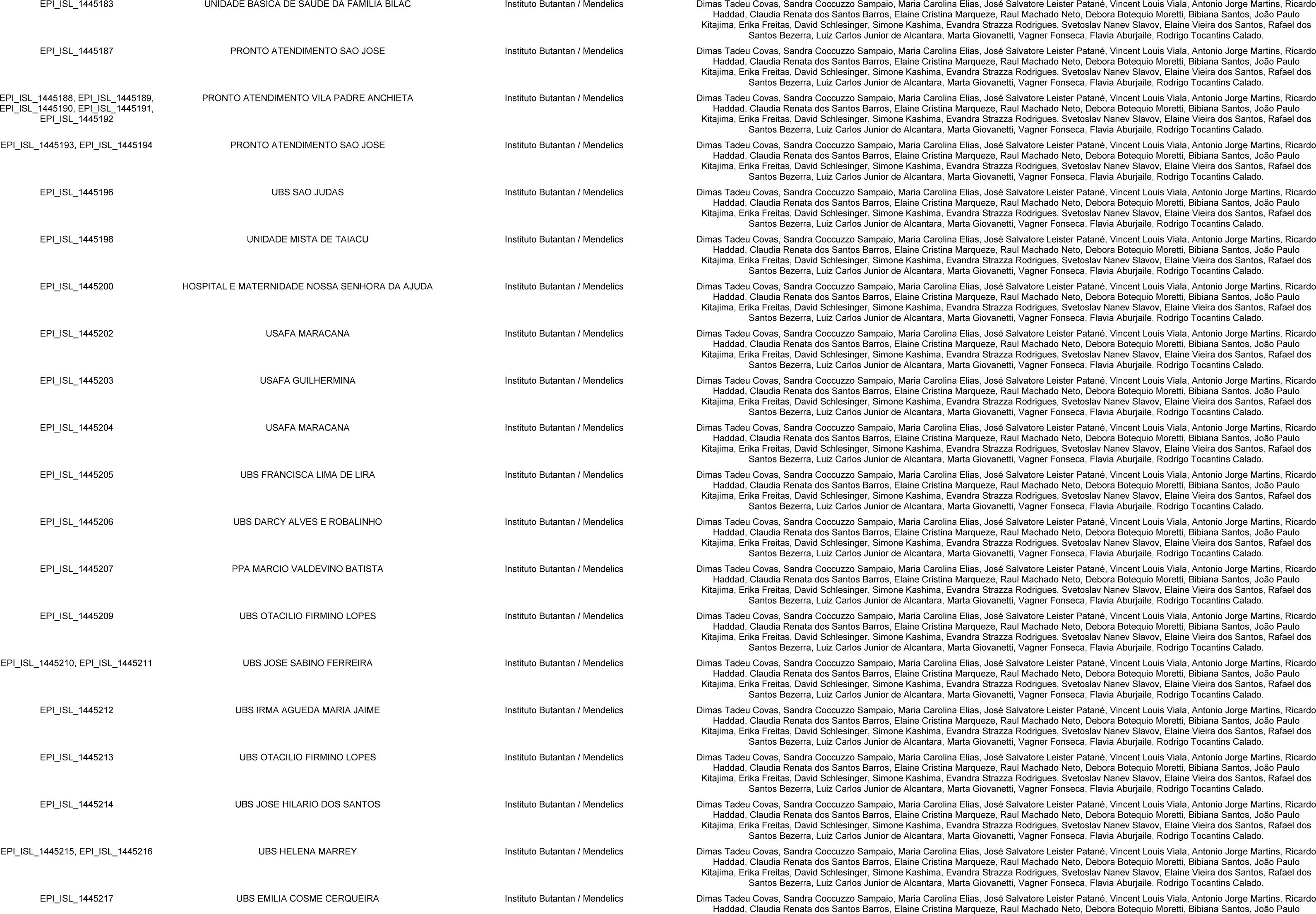
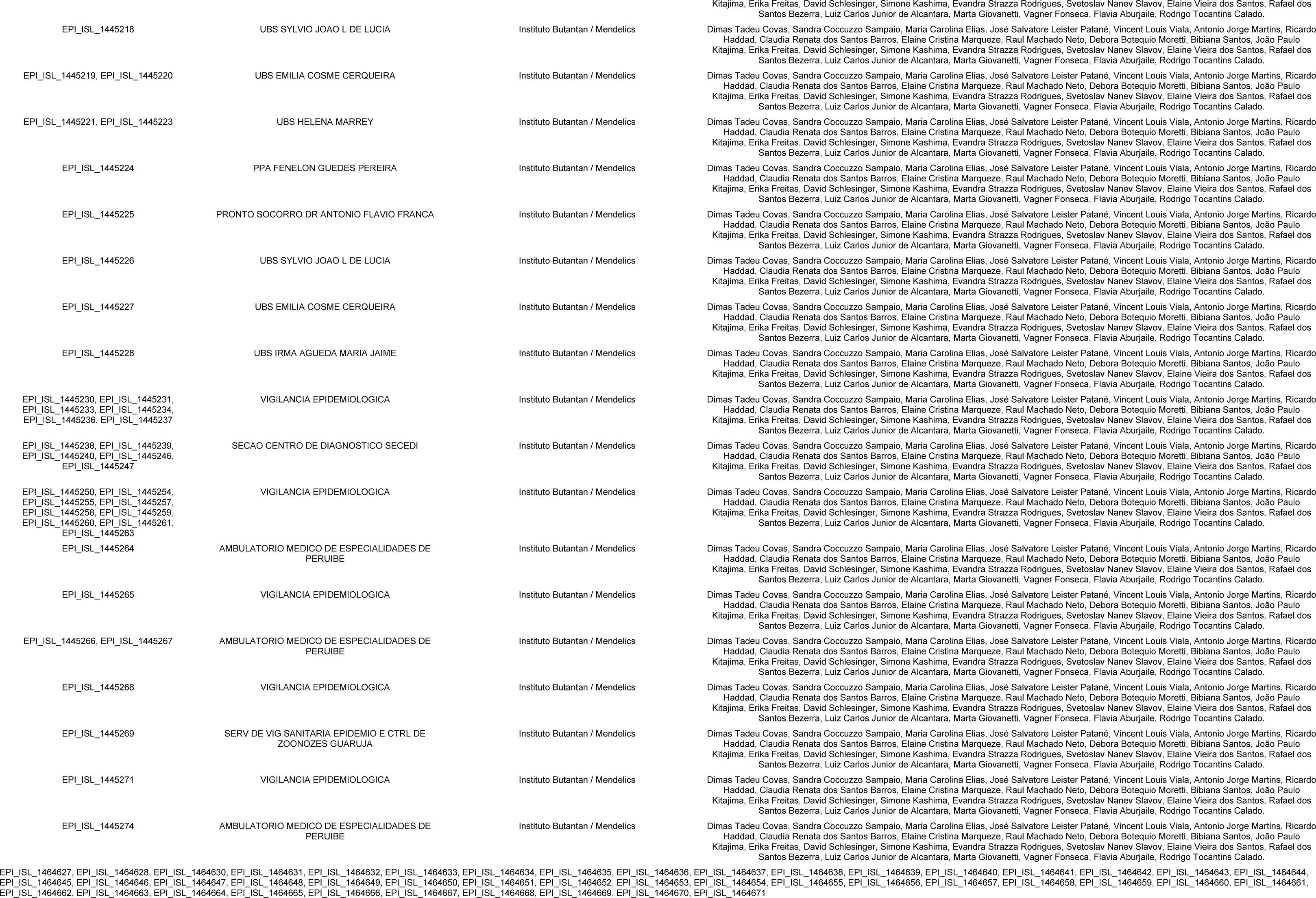
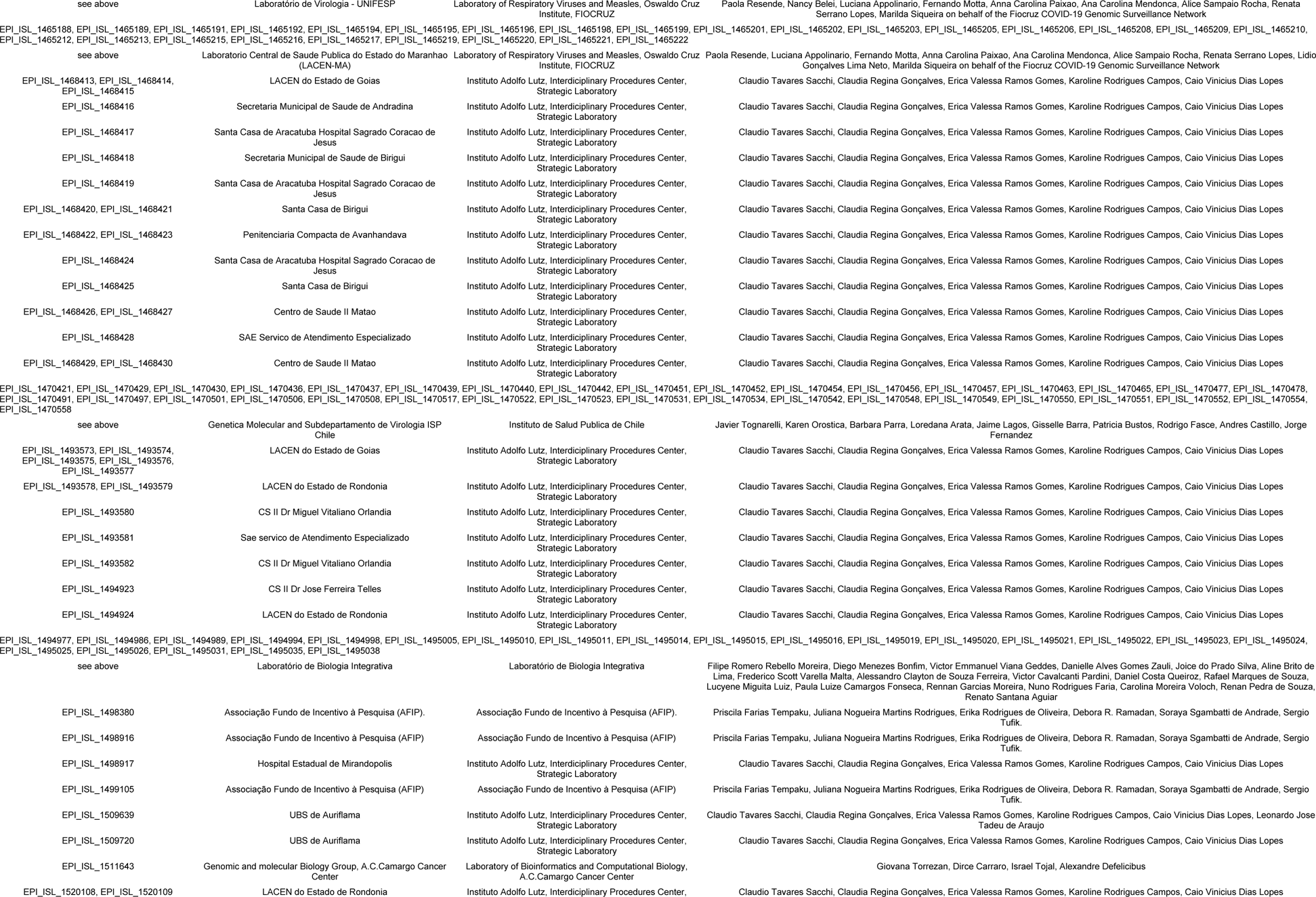
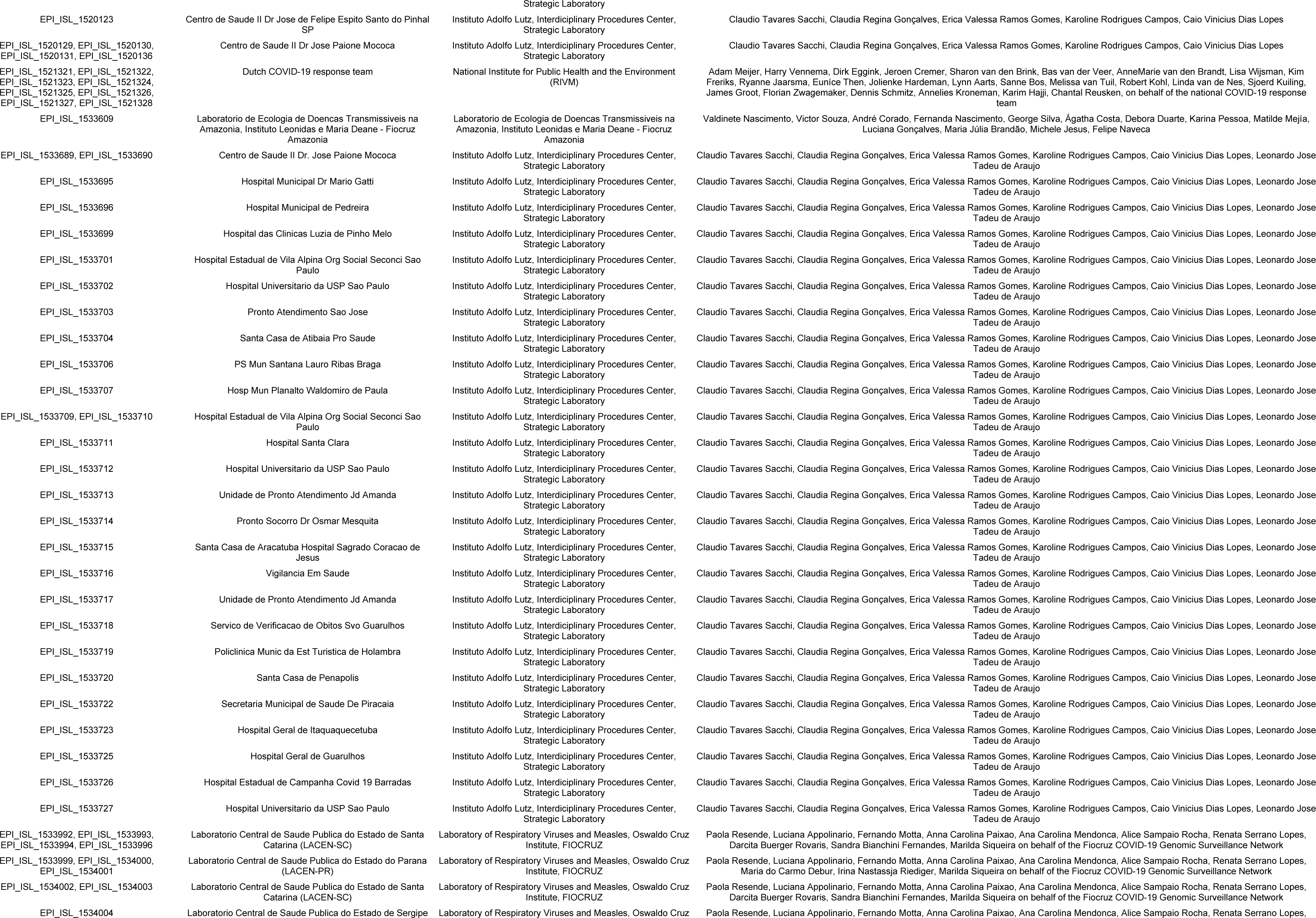
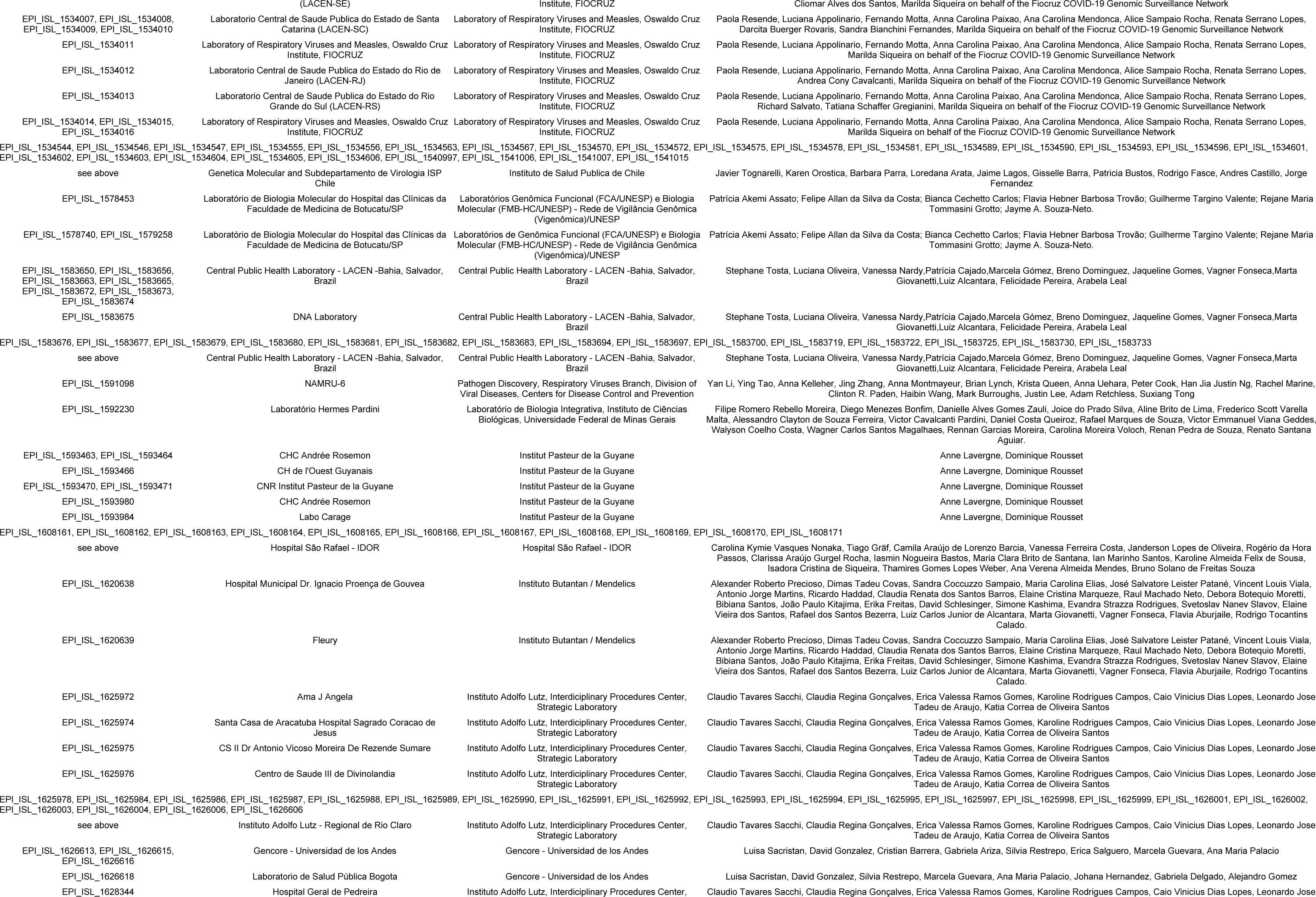
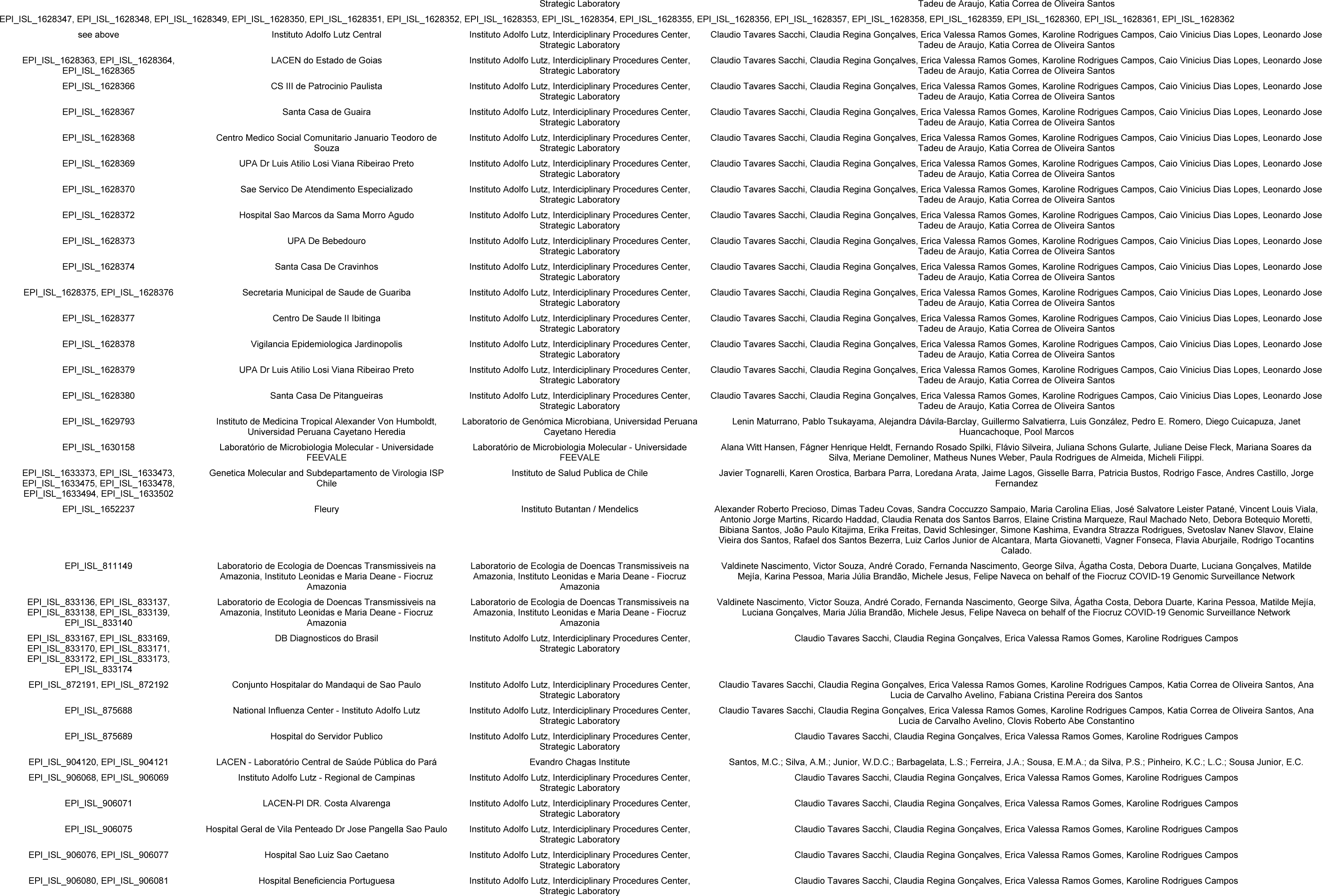
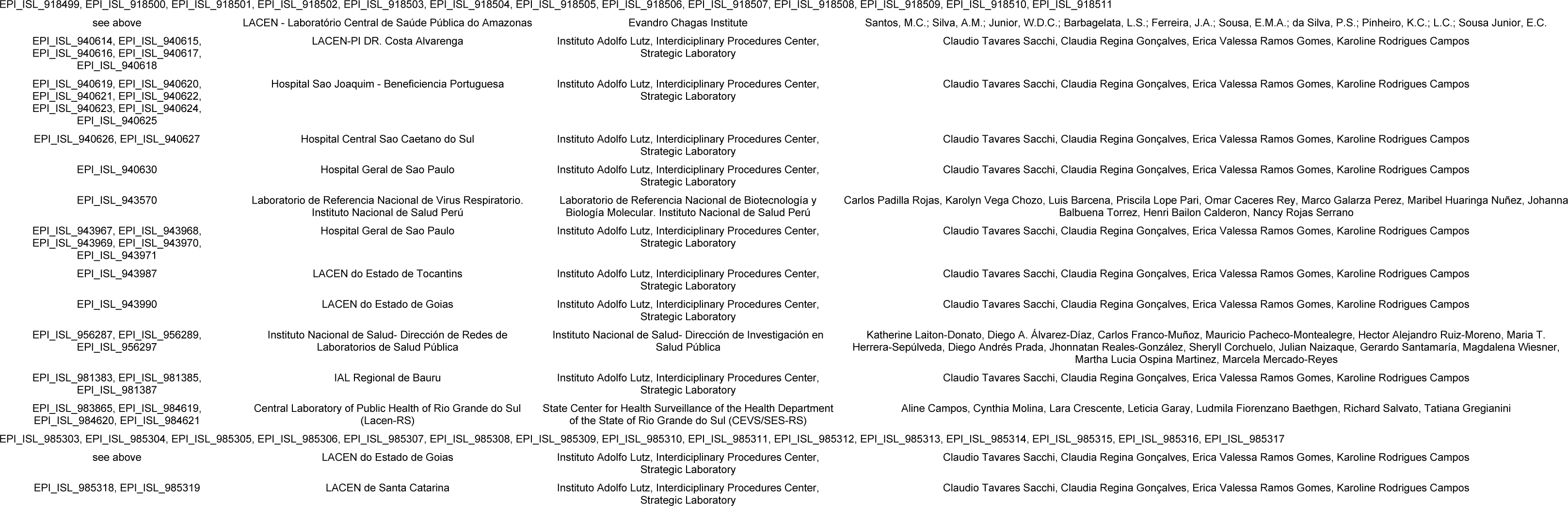

